# Sex differences in the trajectories of plasma biomarkers, brain atrophy, and cognitive decline relative to amyloid onset

**DOI:** 10.1101/2024.09.03.24312664

**Authors:** Cassandra M. Joynes, Murat Bilgel, Yang An, Abhay R. Moghekar, Nicholas J. Ashton, Przemysław R. Kac, Thomas K. Karikari, Kaj Blennow, Henrik Zetterberg, Madhav Thambisetty, Luigi Ferrucci, Susan M. Resnick, Keenan A. Walker

**Author notes:** Corresponding author: Keenan A. Walker, NIH BRC BG RM 04B311, 251 Bayview Blvd, Baltimore, MD 21224. Co-senior authors, equal contribution (Susan M. Resnick, Keenan A. Walker).

## Abstract

**INTRODUCTION:** The factors that influence the progression of Alzheimer’s disease (AD) after individuals become amyloid-positive are poorly understood. This study examines how sex influences the longitudinal trajectories of plasma AD and neurodegenerative biomarkers in the years following a person’s estimated onset of amyloid-β.

**METHODS:** Linear mixed effects modeling investigated overall and sex-specific longitudinal trajectories of plasma biomarkers, brain volumes, and cognition relative to estimated age of amyloid onset in a cohort of 78 amyloid-positive Baltimore Longitudinal Study of Aging participants (n=45 male; follow-up time: 6.8 years [SD 3.31]). Amyloid status was ascertained with ^11^C-Pittsburgh compound B (PiB) PET imaging.

**RESULTS:** After amyloid onset, men displayed steeper increases in pTau181, pTau231, and NfL compared to women. In this same period, men demonstrated steeper declines in brain volume and cognitive performance.

**DISCUSSION:** These findings suggest that sex influences the trajectory of AD pathology, neuronal injury, and symptom progression after individuals become amyloid-positive.

## Introduction

Alzheimer’s disease (AD) is an increasingly common neurodegenerative disease which affects memory and other cognitive domains. In 2021 Alzheimer’s Disease International reported that by 2030 it is estimated a total of 78 million globally will be affected by dementia [1]. Additionally, more women are living with AD than men, nearly 2/3 of those with dementia are female [2]. Despite the difference in AD prevalence, the effect of sex on the trajectory of AD pathological changes and neurodegenerative processes in the preclinical and prodromal stages of AD remains an open question and an active area of investigation [3].

AD is currently defined by the presence of amyloid-β plaques, tau deposition, and clinically assessed dementia [4]. The amyloid cascade hypothesis posits that the accumulation of amyloid-β plaques is the initial step in a chain of pathological processes that lead to AD [5, 6]. However, not all persons who accumulate amyloid-β plaques go on to develop other disease-associated processes, such as tau accumulation and neuroinflammation, which are linked to the expression of clinical symptoms [7, 8]. Moreover, there is a large degree of person-to-person variability both in the age of amyloid onset and the duration between amyloid onset and the expression of mild cognitive impairment (MCI) or dementia [9–12]. Determining the demographic and biological factors that influence the rate of downstream disease progression following the emergence of amyloid-β plaques is essential for advancing our understanding of AD biology and refining the amyloid cascade hypothesis.

Although PET imaging remains the gold standard for the *in vivo* identification of amyloid-β plaques and tau burden, circulating peripheral proteins associated with AD pathology (namely the ratio of amyloid-β_42_ to amyloid-β_40_ [Aβ_42_/Aβ_40_], and levels of phosphorylated tau [pTau]), neuronal injury (neurofilament light [NfL]), and a type of astrocyte-mediated inflammation (reactive astrocytosis; glial fibrillary acidic protein [GFAP]) can now be quantified with minimally invasive plasma-based measures [13]. Additionally, previous work by our lab and others has shown that accumulation of cortical amyloid-β follows a consistent trajectory after onset, allowing age of amyloid onset to be estimated in those with an amyloid-positive PET scan [9, 10, 12, 14]. This ability to estimate when a person began accumulating amyloid is particularly relevant to understanding sex differences in AD because there is evidence that women begin accumulating amyloid earlier than men [15, 16]. Thus, the examination of biomarker trajectories in the years following estimated age of amyloid onset could be used to gain insight into the patterns and determinants of the neurobiological changes that may drive clinical progression following amyloid deposition. Ultimately, an understanding of biomarker changes with respect to the accumulation of amyloid plaques will allow a better characterization of the preclinical and prodromal phase of AD.

To further advance our understanding of the sequence of pathophysiological processes that occur in the preclinical and prodromal phase of AD, the present study used data from the Baltimore Longitudinal Study of Aging (BLSA) to examine the trajectories of plasma biomarkers of AD pathology (Aβ_42_/Aβ_40_, pTau181, and pTau231), neuronal injury (NfL), and reactive astrogliosis (GFAP) after estimated age of amyloid onset. Additionally, we determined whether these plasma biomarker trajectories differed by sex and subsequently examined whether the sex differences extended to brain atrophy and cognitive decline. Previous findings from the BLSA have shown amyloid positive men have steeper rates of volumetric declines in the entorhinal cortex and parahippocampal gyrus [17], as well as steeper declines in performance on several cognitive tests[18]. In light of these findings, we hypothesized that men would have steeper accumulation of AD and neuronal injury plasma biomarkers, and more rapid declines in brain volume and cognition following amyloid onset.

## Methods

### 2.1 Participants

The Baltimore Longitudinal Study of Aging (BLSA) is a community-based study which follows participants over their lifespan. BLSA visits occur every 4 years for participants less than age 60, every 2 years between ages 60 and 79, and annually for participants aged 80 or older. Participants in the PET neuroimaging sub-study of the BLSA are age 55 and older. All participants in the current study had amyloid PET imaging collected, after QC of the amyloid PET data, cortical amyloid levels were quantified for 225 participants. The present study focuses on any BLSA participants who had a positive amyloid PET scan at one or more of their PET visits. For detailed information about participant inclusion and exclusion see **sTable1**, **sTable3**, and **sFig 1**. This study was approved by the local institutional review boards overseeing the BLSA and the PET imaging studies, and participants provided written informed consent at each study visit.

### 2.2 Amyloid PET imaging and estimation of amyloid onset age

PET amyloid imaging studies were conducted at the Johns Hopkins PET Facility, using ^11^C-Pittsburgh Compound B (PiB) and either a GE Advance or a Siemens High Resolution Research Tomograph (HRRT) scanner. Procedures for image acquisition and analysis, including harmonization between scanners are detailed in [19]. Briefly, approximately of 555 MBq of PiB was injected as a bolus, and 70-minute dynamic scans were acquired. Scans were reconstructed into 33 timeframes using filtered back projection (GE Advance) or ordered subset expectation maximization (Siemens HRRT). Distribution volume ratio (DVR) images were computed using a spatially constrained simplified reference tissue model with cerebellar gray matter as the reference region [20]. Mean cortical amyloid burden was calculated as the average of the DVR values in the cingulate, frontal, parietal, lateral temporal, and lateral occipital regions, excluding the pre- and post-central gyri. Following harmonization of mean cortical amyloid burden between scanners, amyloid positivity was determined using a 2-class Gaussian mixture model fitted to baseline amyloid values. The threshold for amyloid positivity was a DVR of 1.065.

After its onset, amyloid accumulation progresses at a consistent rate across individuals. Once a participant has an amyloid positive scan it is possible to estimate the age at which they first became positive by longitudinally extrapolating from the level of cortical amyloid observed on their scan(s) until minimal amyloid levels are reached. Age at amyloid onset was estimated using a piecewise linear mixed effects model, as described by Bilgel et al. [12]. There was no observed difference in estimated age of amyloid onset between men and women, as described in [10] and shown in **Table 1** and **sFig 4**.

**Table 1.**
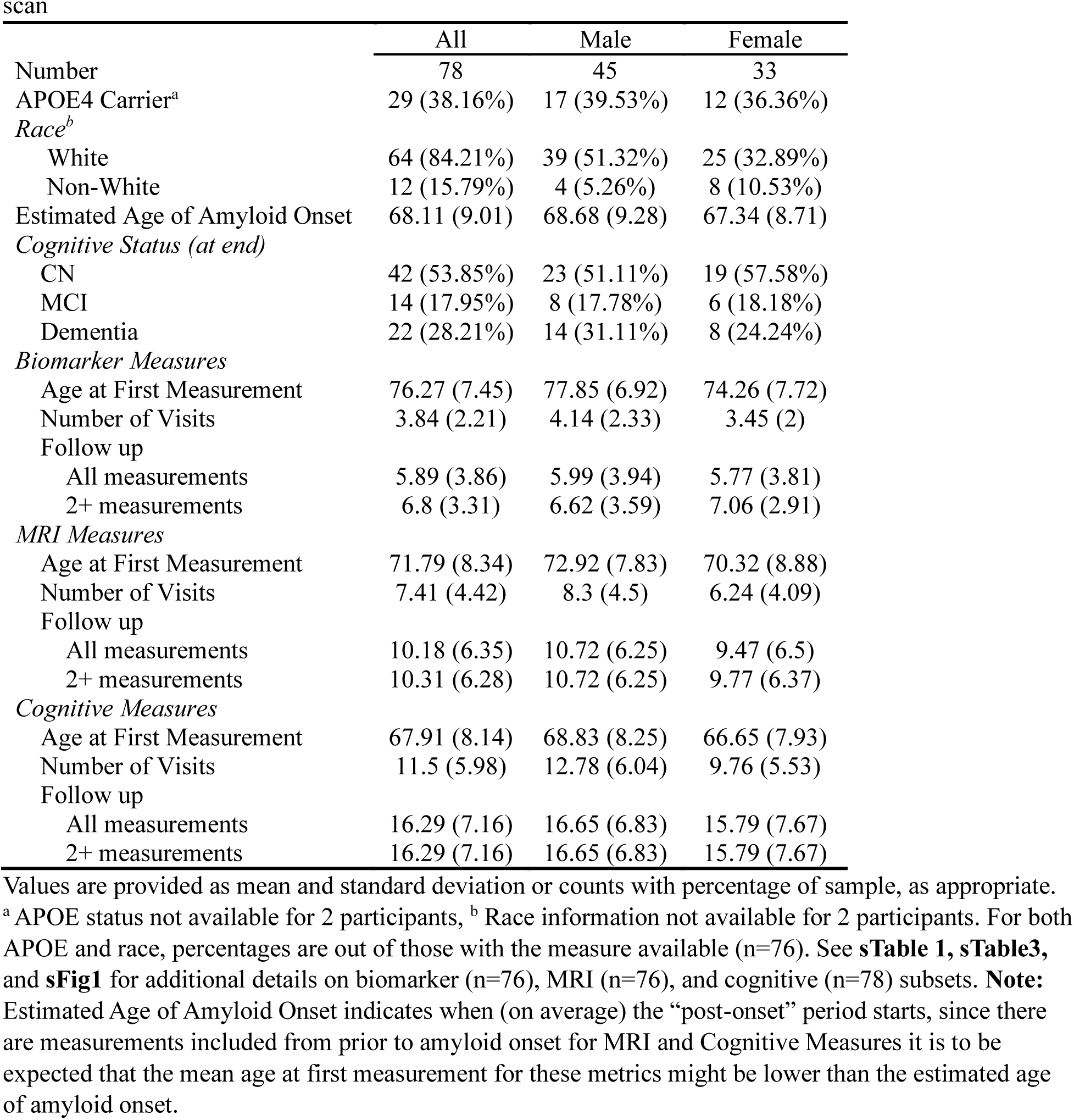
Demographic information for BLSA participants confirmed to be amyloid positive on PiB PET scan.

### 2.3 Plasma biomarkers

Blood was drawn at the same visit as PET and MRI scans, centrifuged, and aliquoted plasma was stored at -80°C until analysis. Aβ_40_, Aβ_42_, NfL, and GFAP assays were performed using the Single molecule array (Simoa) Neurology 4-Plex E assay on a Simoa HD-X Analyzer (Quanterix Corporation). Protein levels were assessed in duplicate and the mean of the two measurements was used here. The intraassay coefficients of variation were estimated to be 1.5 for Aβ_40_, 1 for Aβ_42_, 4.9 for GFAP, and 4.8 for NfL. pTau levels were assessed by investigators in the Clinical Neurochemistry Laboratory at the University of Gothenburg using in-house assays on a Quanterix Simoa HD-X Analyzer [21, 22]. Repeatability coefficients were 5.1% and 5.5% for the pTau181 assay at concentrations of 11.6 and 15.5 pg/mL, respectively. Repeatability coefficients were 3.4% and 7.4% for the pTau231 assay at concentrations of 31.6 and 42.7 pg/mL, respectively. Estimated glomerular filtration rate (eGFR) was calculated using measured levels of serum creatinine following the 2009 CKD-epi equation [23].

### 2.4 MR imaging

Anatomical MR scans were acquired on GE Signa 1.5 T and Philips Achieva 3 T scanners, using the SPGR and MPRAGE sequences respectively. MRI measures assessed in this study were total brain volume (TBV), total grey matter (GM) volume, total white matter (WM) volume, ventricular volume, two composite regions of interest (ROIs), and two imaging-based scores (Spatial Patterns of Abnormality for Recognition of Early AD [SPARE-AD] [24] and Brain Predicted Age Difference [Brain-PAD] [25]). All regions for the MRI measures were segmented and labeled using the MUlti-atlas region Segmentation utilizing Ensembles (MUSE) software package [26]. Volumes were harmonized across scanners [27, 28] and adjusted for each participant’s total intracranial volume [29]. The AD Signature ROI was comprised of (bilateral) volumes of the hippocampus, entorhinal cortex (ERC), parahippocampal gyrus (PHG), precuneus, and posterior cingulate gyrus. The medial temporal lobe (MTL) ROI consisted of the hippocampus, ERC, PHG, and amygdala.

Calculations of SPARE-AD and SPARE-Brain Age (SPARE-BA) scores from anatomical MRI scans have been described previously [24, 25, 30]. In brief, for SPARE-AD a support vector machine (SVM) was trained on a large number of anatomical scans from those clinically diagnosed with AD as well as from AD-free controls to estimate the degree to which each scan resembles the brain of someone with AD. Thus, SPARE-AD is a measure of how “AD-like” a given MR image is, with higher scores indicating a brain that looks more similar to the brain of someone with AD. SPARE-BA, which estimates the age that the person was at the time of the scan, was calculated similarly, however only images from healthy control participants were used for training the SVM. Brain-PAD was calculated by subtracting the participant’s actual age at the time of scan from their SPARE-BA estimated brain age. Thus, having a positive Brain-PAD score means that the brain age estimated by SPARE-BA is older than the person’s chronological age (i.e., the brain looks older than it should). A positive longitudinal slope on Brain-PAD means that the participant’s estimated brain age is increasing faster than their chronological age (i.e. they are experiencing accelerated brain aging).

### 2.5 Cognitive assessment

Cognitive assessments were administered by trained testers at each BLSA visit and evaluated a wide array of cognitive abilities including memory, executive function, and visuospatial abilities. Longitudinal cognitive measures included in this study are the California Verbal Learning Test (CVLT) – list learning/immediate free recall and long delay free recall; the Mini Mental State Examination (MMSE); the difference in time to complete Trail Making Test A and Trail Making Test B (Trail Making Test A-B; an indicator of executive function, with a larger difference in completion times indicating lower executive function); the Digit Symbol Substitution Test (DSST), a measure of executive function and processing speed; and the Card Rotations Test, a measure of spatial rotational ability. All cognitive measures, except the Trail Making Test A-B, are reported as scores where a higher score indicates better performance/less cognitive decline. As mentioned above, the Trail Making Test A-B is the subtraction of the time taken to complete two different tasks, and a larger gap in these scores indicates poorer executive function.

### 2.6 Statistical analysis

In this study we characterized sex differences in trajectories of biomarker, MRI, and cognitive measures relative to each participant’s estimated age of amyloid onset. We used linear mixed effects (LME) models to estimate longitudinal trajectories for the measures of interest using the following predictors: estimated age of amyloid onset, years since estimated amyloid onset (henceforth “time”), sex, and the interaction of sex and time (sex × time). For all analyses except those including amyloid negative participants (i.e. **Section 3.1**), we anchored the time term at age of amyloid onset rather than using age directly to allow us to investigate amyloid based changes in trajectories independent of the age at which a person first became positive for cortical amyloid. We first examined the trajectories of plasma biomarker, brain volume, and cognitive measures relative only to amyloid onset using LME models which included all the aforementioned covariates except the sex × time interaction term. LMEs for the plasma biomarkers also included estimated glomerular filtration rate (eGFR) as a covariate, to account for differential clearing of the proteins from the blood by the kidneys. After characterizing the general longitudinal trajectories of our measures of interest, we fit a second set of LME models (primary analysis) with a sex × time interaction term to characterize how sex influences longitudinal trajectories of each outcome measure. Since our focus is on the fixed effects part of the LME models, we included only a random intercept to account for the within subjects correlation of the data for our LME models, as inclusion of a random slope caused convergence issues for some of the LME models. For MRI and cognitive measures, we had sufficient data points to allow for separate LME models to be fit both before and after amyloid onset; however, our focus (defined *a priori*) was on the post-amyloid onset results, which offered greater statistical power and represent a phase in the disease course that is highly relevant to the progression from asymptomatic to symptomatic AD. All analyses were performed in R (version 4.4.1), with LME analysis performed using the lmer function from the lmerTest package [31, 32].

## Results

### 3.1 Cortical amyloid influences trajectory of ADRD biomarkers in plasma

We first determined whether eventual amyloid-β status, as defined by ever having an amyloid positive PiB PET scan, modified the slope of our biomarkers of interest in 221 participants with available data (age at first biomarker measurement: 73.07 [8.03 SD]; 53.1% women; 22.2% non-white). It is not possible to estimate an age of amyloid onset in amyloid negative individuals, thus for this analysis the baseline visit was defined as the first visit for which a person had plasma collected. This time scale is distinct from the baseline time (i.e. estimated age of amyloid onset) used for all subsequent analyses. As illustrated in **Figure 2**, greater increases in biomarker level were observed for pTau181, pTau231, and GFAP among amyloid-positive participants, whereas steeper decreases in Aβ_42_/Aβ_40_ – a pattern consistent with greater amyloid accumulation – was observed among amyloid-negative participants (**sTable 2**).

**Figure 1:**
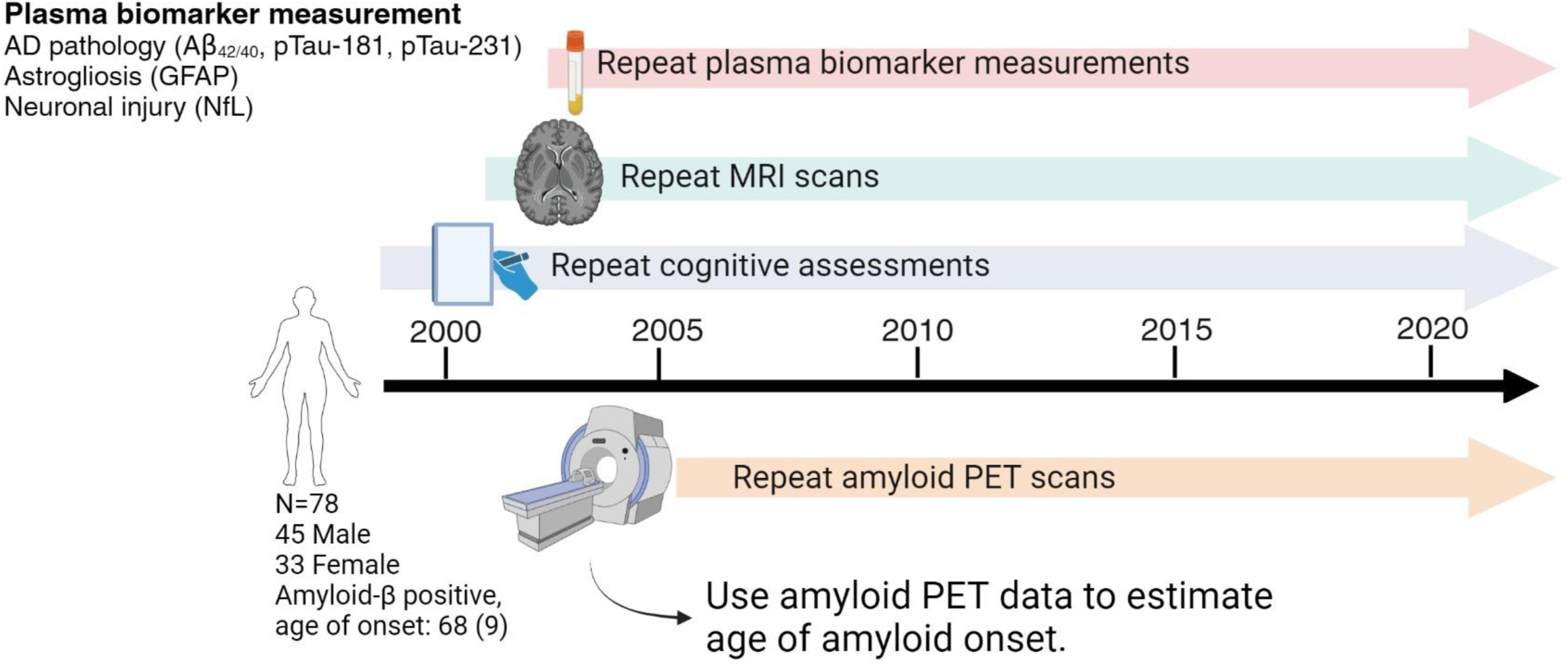
Summary of data collection/study design. The Baltimore Longitudinal Study of Aging began in 1958 and has a rolling study design, wherein participants are continuously recruited, therefore each participant enters at a different time. After enrollment, longitudinal study visits occur every 1, 2, or 4 years depending on a person’s age. Collection of MRIs and cognitive assessments began in 1994; Pittsburgh compound B (PiB) PET scanning began in 2005. Plasma was collected at each baseline 3 T MRI visit and at all PiB PET visits, and frozen until analysis. Figure created with biorender.com.

**Figure 2:**
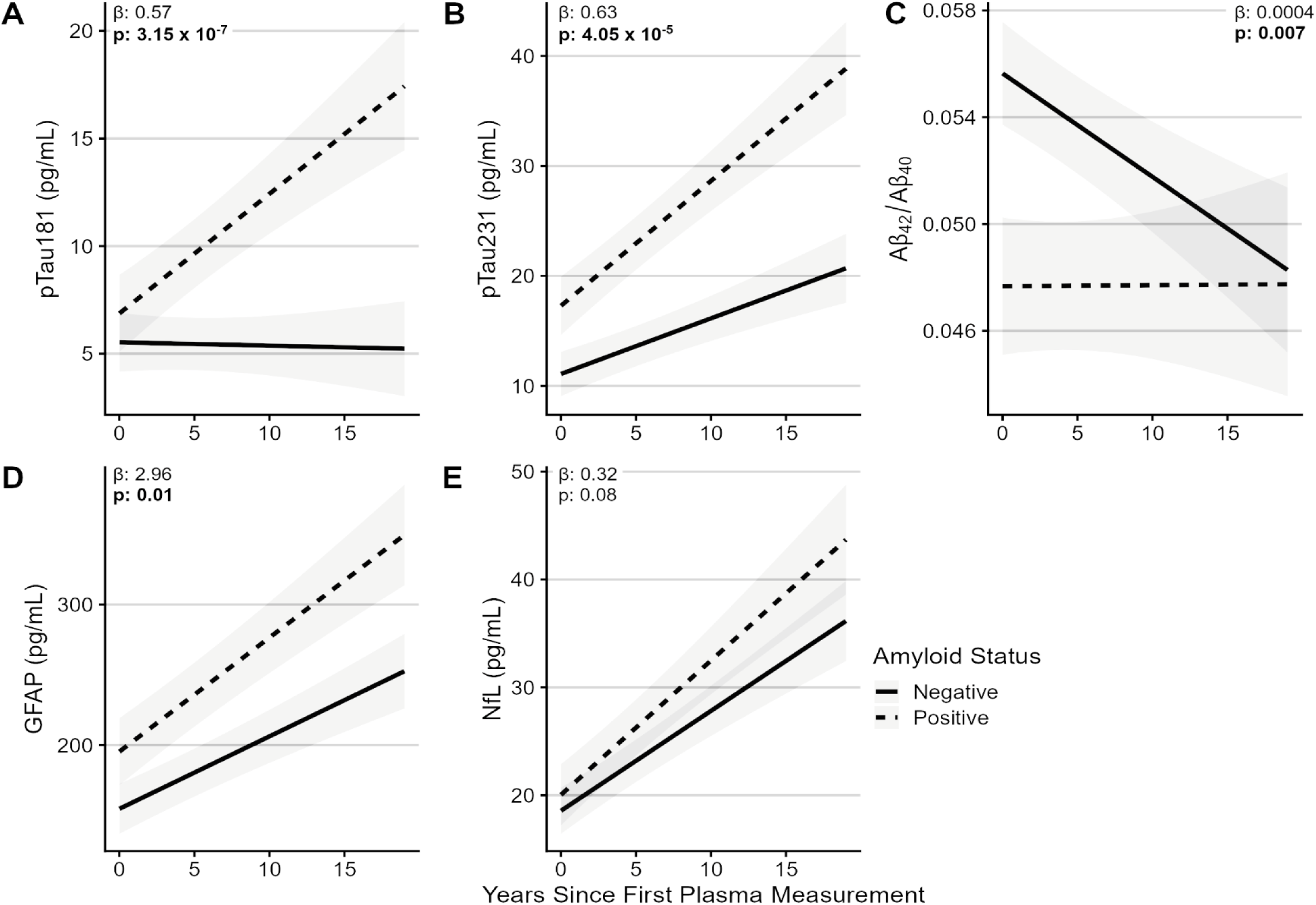
Biomarker trajectories relative to years since first biomarker measurement, stratified by amyloid status at latest observation. **A-E.** The amyloid-specific trajectories of each biomarker as estimated by a linear mixed effects model. β estimates and p-values listed on each panel are for the amyloid status × time interaction term in each linear mixed effect model. For visualization purposes, lines have the centered age at first biomarker visit set equal to zero (i.e., lines plotted show accumulation rates for a person having an age at first biomarker visit equal to the mean age of first biomarker visit). **Abbreviations:** Aβ – Amyloid-β, GFAP – glial fibrillary acidic protein, NfL – neurofilament light, pTau – phosphorylated Tau

### 3.2 ADRD plasma biomarker changes following the onset of cortical amyloid

After demonstrating that the presence of cortical amyloid-β modified the trajectory of ADRD biomarkers in plasma, we characterized the longitudinal trajectories of 5 plasma biomarkers (Aβ_42_/Aβ_40_, pTau181, pTau231, GFAP, and NfL) after estimated age of amyloid onset in 75 amyloid-positive participants (age at first biomarker measurement: 76.27 [7.45 SD]; 44.0% women; 16.0% non-white; **Table 1**, **sTable 3**).

Following estimated amyloid onset, we observed significant longitudinal increases in pTau181 (0.06 standardized units [s.u]/yr), pTau231 (0.08 s.u./yr), GFAP (0.06 s.u./yr), and NfL (0.07 s.u. /yr), and a non-significant longitudinal decrease in Aβ_42_/Aβ_40_ (-0.004 s.u./yr) (**Fig. 3A**, **sTable 4**). An examination of the correlations between individuals’ rates of change in biomarkers found that the rates of change in pTau181 and pTau231 were strongly correlated (r = 0.6, p = 9.39E-9), and that rate of NfL change was modestly correlated with rate of change in pTau181, pTau231, and GFAP (r = 0.26, 0.44, and 0.4 respectively, p’s = 0.02, 7.89E-5, and 0.0004; **sFig. 2A**).

**Figure 3.**
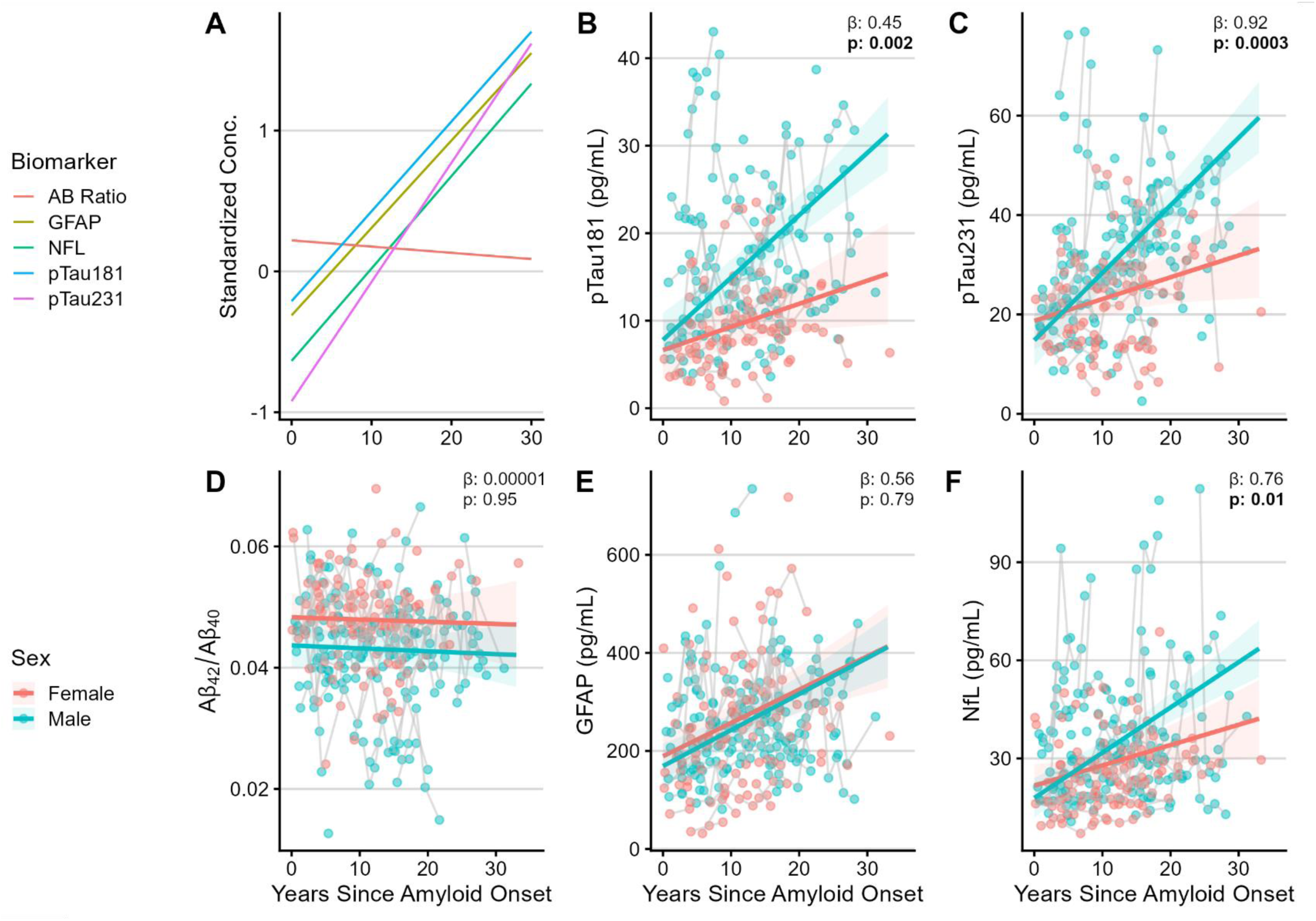
Combined and sex-specific longitudinal trajectories of plasma biomarkers of Alzheimer’s disease and neurodegeneration. **A**. The standardized (z-scored) concentrations of each biomarker, once observations have been centered based on the participant’s estimated age of amyloid onset. **B-F.** The sex-specific trajectories of each biomarker as estimated by a linear mixed effects (LME) model overlaid on the observed biomarker concentration data. Each point represents a single observation of the biomarker’s concentration, with grey lines connecting repeated observations from the same participant. β estimates and p-values listed on each panel are for the sex × time interaction term in each LME model. The reference group for the interaction terms is women. Thus, a positive β value for the interaction term indicates that men have more/steeper longitudinal increase in that metric than women. For visualization purposes, the colored lines showing the LME model have all other covariates set to the mean of the sample (for instance, lines plotted show accumulation rates for a person having an age of amyloid onset equal to the mean age of amyloid onset). For the z-scored lines (**A**), sex and eGFR were included as covariates and the lines plotted are the trajectories for females (the reference group). **Abbreviations:** Aβ – Amyloid-β, GFAP – glial fibrillary acidic protein, NfL – neurofilament light, pTau – phosphorylated Tau

In the primary analyses examining sex differences in rates of change in the plasma biomarkers, we found significant sex differences in longitudinal accumulation of pTau181 (**Fig. 3B**, interaction β = 0.45 ± 0.15, p = 0.003), pTau231 (**Fig. 3C**, interaction β = 0.92 ± 0.25, p = 0.0003), and NfL (**Fig. 3F**, interaction β = 0.76 ± 0.30, p=0.01) following amyloid onset. Men had faster accumulation of pTau181, pTau231, and NfL than women following amyloid onset. Notably, correlations between rates of change in plasma biomarkers also differed (qualitatively) between men and women (**sFig. 2B-D**).

### 3.3 Brain volume changes relative to the onset of cortical amyloid

We next investigated whether the sex difference observed for longitudinal change in pTau and NfL levels relative to amyloid onset would also be observed for MRI measures of brain volume changes. Since there were sufficient pre-onset observations, LME models were fit separately to data from before and after cortical amyloid onset, to determine whether any observed post-onset trajectory changes were also occurring prior to onset of PET-observable amyloid.

This analysis included 76 participants (age at first MRI measurement: 71.79 [8.34 SD]; 43.4% women; 16.2% non-white; **Table 1**, **sTable 3**), 23 of whom were included in the pre-amyloid onset analyses. Total brain, grey matter, and white matter volumes showed significant longitudinal decreases both before and after estimated age of amyloid onset, as did the volumes for MTL and AD Signature ROIs. Similarly, ventricular volumes, as well as SPARE-AD and Brain-PAD scores, showed significant longitudinal increases before and after estimated age of amyloid onset (**sTable 5**).

An analysis of sex-specific longitudinal trajectories of these measures found that, relative to women, men had significantly steeper declines after estimated amyloid onset in total brain (interaction β = -816.4 ± 264.12, p=0.002) and grey matter (interaction β = -747.01 ± 198.84, p=0.0002) volumes, and a faster increase in ventricular size (interaction β = 336.57 ± 78.24, p = 0.00002). Additionally, men had steeper increases than women in SPARE-AD (interaction β = 0.04± 0.008, p = 0.0000002) and Brain-PAD (interaction β = 0.22 ± 0.06, p = 0.0005) scores, suggesting greater AD-like patterns of atrophy and accelerated brain aging, respectively, among men following amyloid onset. Examination of MRI measures prior to amyloid onset showed that all metrics investigated except SPARE-AD show significant sex × time interactions consistent with the post-onset findings (**Fig. 4**, **sTable 6**), suggesting that – with the exception of the measure of AD-relevant atrophy (SPARE-AD) – sex differences in rates of brain atrophy might begin before the onset of PET detectable amyloid. Individual analyses of the regions included in the composite ROIs, MTL and AD Signature, can be found in Supplementary Table 9 and visualized in Supplementary Figure 3.

**Figure 4:**
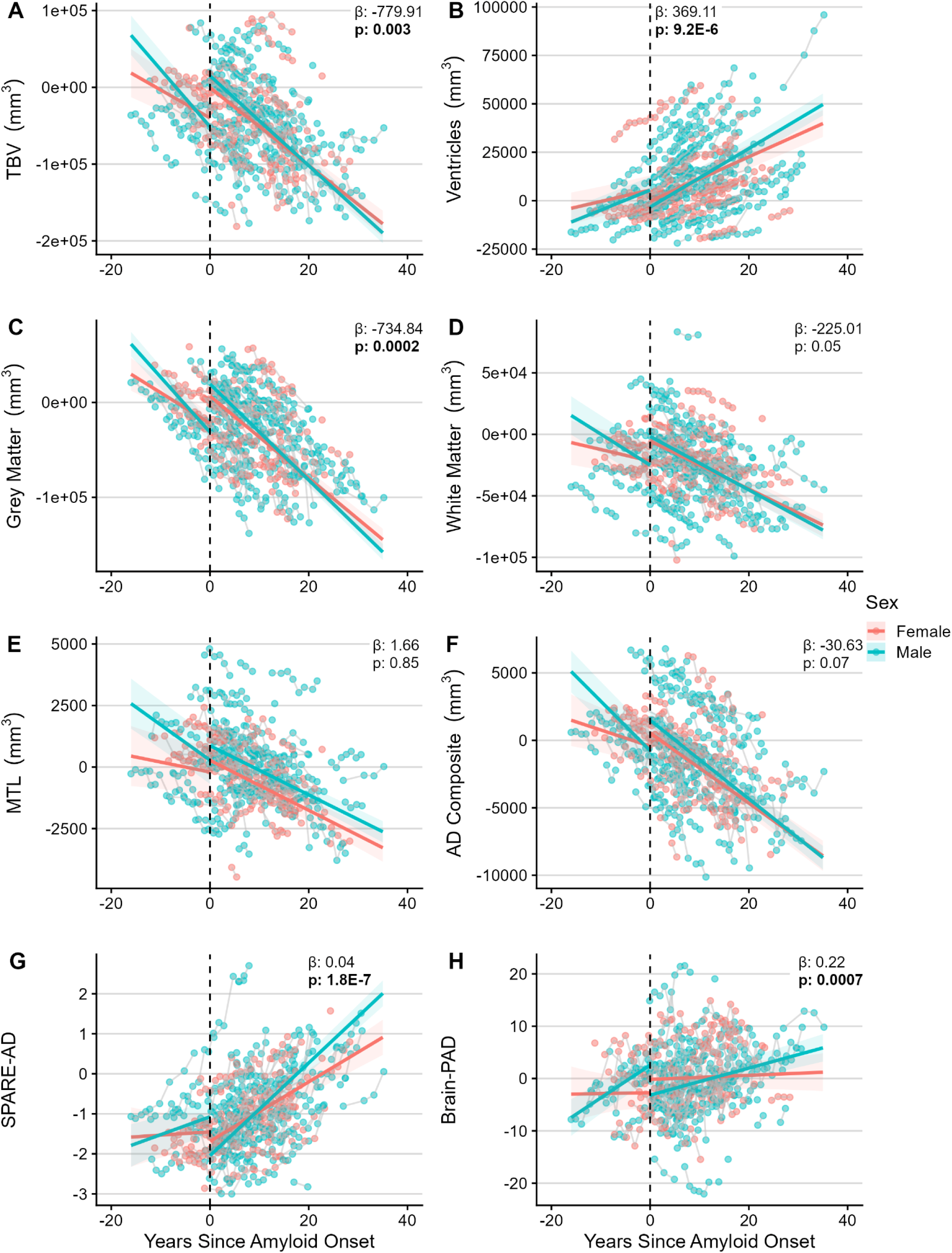
Sex-specific trajectories of MRI based measures before and after amyloid onset. Sex specific trajectories as estimated by linear mixed effects models for MRI-defined brain volumes of (**A**) total brain, (**B**) total ventricular size, (**C**) total grey matter, (**D**) total white matter, (**E**) medial temporal lobe, and (**F**) AD composite region. Sex specific trajectories as estimated by linear mixed effects models for scores of patterns of brain changes indicating (**G**) Alzheimer’s-like-ness and (**H**) estimated brain age derived from a structural MRI scan. Each point represents a single observation of the ICV adjusted and harmonized MRI measure (meaning each point represents the difference in volume from expected, given the person’s ICV), with grey lines connecting repeated observations from the same participant. β estimates and p-values listed on each panel are for the sex × time interaction term in each LME model. The reference group for the interaction terms is women. Thus, a negative β value for the interaction term indicates that men have more/steeper longitudinal decline in that metric than women. For visualization purposes, the colored lines showing the LME model have all other covariates set to the mean of the sample (for instance, lines plotted show accumulation rates for a person having an age of amyloid onset equal to the mean age of amyloid onset). **Abbreviations:** Brain PAD – brain predicted age difference; ICV – intracranial volume; MTL – medial temporal lobe; SPARE-AD – spatial pattern of abnormality for recognition of early Alzheimer’s disease; TBV – total brain volume

### 3.4 Cognitive changes relative to the onset of cortical amyloid

Lastly, we investigated sex differences in changes in cognitive measures relative to amyloid onset. Given the consistent findings for biomarkers and imaging measures, as well as previous BLSA findings [18], we anticipated men would have steeper cognitive decline than women after amyloid onset.

The analysis of cognitive performance following the estimated onset of cortical amyloid included 78 participants (age of first cognitive assessment: 67.91 [8.14 SD]; 42.3% women; 15.8% non-white; **Table 1**, **sTable 3**), 43 of whom were included in the pre-amyloid onset analyses. Performance on the MMSE and on measures of processing speed and executive function (Trail Making Test A-B, DSST), memory (CVLT), and visuospatial abilities (Card Rotations Test) declined significantly following the estimated onset of cortical amyloid, whereas only performance on measures of processing speed and executive function showed significant declines prior to the estimated age of amyloid onset (**sTable 7**).

As shown in **Fig. 5**, cognitive trajectories after amyloid onset differed by sex for the Card Rotations Test (interaction β = -0.75 ± .24, p = 0.00052; **sTable 8**, **Fig. 5F**) and for the Trail Making Test A-B (interaction β = -0.86 ± 0.43, p = 0.048; **sTable 8**, **Fig. 5B**), indicating men have significantly steeper declines than women on these measures of visuospatial and executive abilities. Preceding amyloid onset, we found steeper declines in Trail Making Test A-B performance among women compared to men, in contrast to what is observed following estimated age of amyloid onset (**sTable 8, Fig. 5B**).

**Figure 5:**
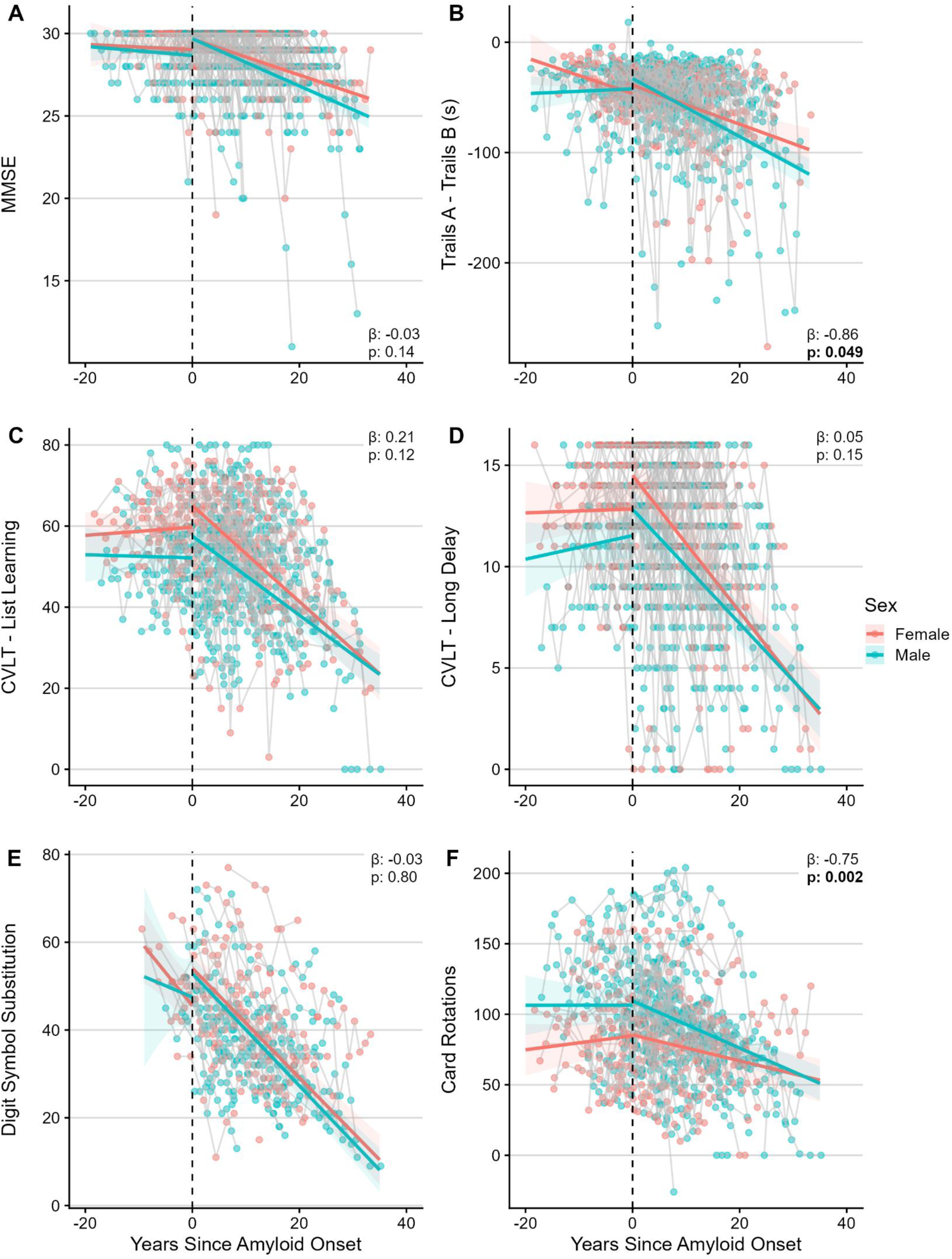
Sex-specific longitudinal trajectories of cognitive measures before and after amyloid onset. Sex specific cognitive trajectories as estimated by linear mixed effects models for the (**A**) MMSE, (**B**) Trail Making Test, (**C**) CVLT List-Learning, (**D**) CVLT Long Delay, (**E**) Digit Symbol Substitution, and (**F**) Card Rotations. Each point represents a single observation of cognitive performance, with grey lines connecting repeated observations from the same participant. β estimates and p-values listed on each panel are for the sex × time interaction term in each LME model. The reference group for the interaction terms is women. Thus, a negative β value for the interaction term indicates that men have more/steeper longitudinal decline in that metric than women. For visualization purposes, the colored lines showing the LME model have all other covariates set to the mean of the sample (for instance, lines plotted show accumulation rates for a person having an age of amyloid onset equal to the mean age of amyloid onset). **Note:** All measures except Trail Making Test are provided as scores where a higher score indicates higher cognitive function, the Trail Making Test is a difference in time to completion for Trail Making Test A and Trail Making Test B thus reported in seconds, a larger difference in the two scores (i.e. a more negative score) indicates reduced cognitive performance. **Abbreviations:** CVLT – California Verbal Learning Test, MMSE – mini mental state examination.

Given the sex differences observed in the longitudinal trajectories of pTau, brain volumes, and cognition after the estimated onset of cortical amyloid, we next characterized the duration between amyloid onset and the first diagnosis of (1) any form of cognitive impairment (including both MCI and dementia) and (2) dementia only. We find that men have a qualitatively shorter time to the onset of cognitive impairment (median 14.85 years; IQR: 6.78, 20.17; **Fig. 6A**) and dementia (median 15.55 years; IQR: 8.35, 21.1; **Fig. 6B**) after estimated amyloid positivity, relative to women who show a median duration of 15.95 years (IQR 10.75, 22.1) and 20.85 years (IQR 16.37, 25.75) to onset of cognitive impairment and dementia, respectively.

**Figure 6:**
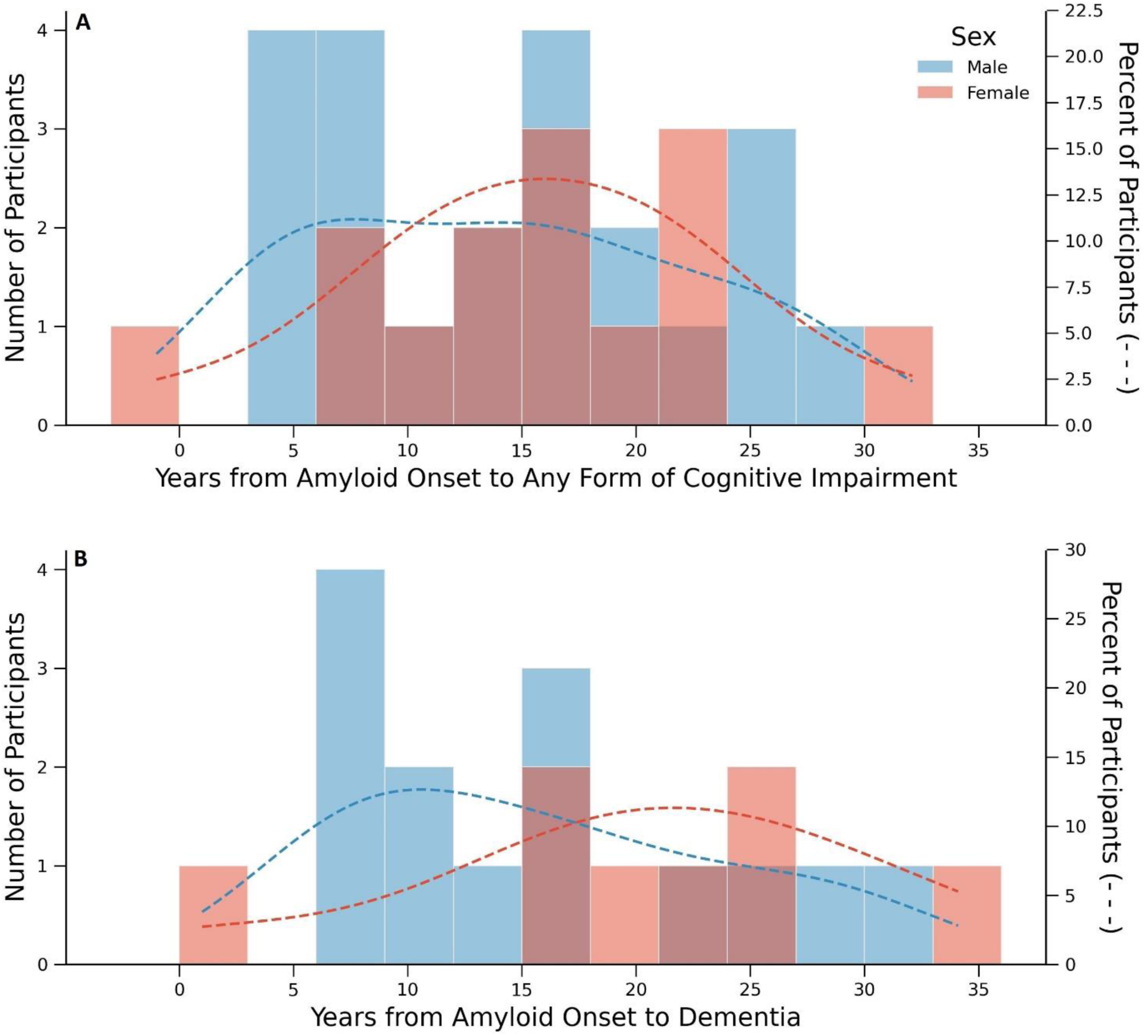
Sex-specific distributions of time between amyloid onset and cognitive impairment. Panel A reflects time to MCI diagnosis for 32 of the 36 participants (20 men, 12 women). The other 4 participants had a diagnosis of dementia without a prior diagnosis of MCI (2 men, 2 women) and thus their time to dementia onset was used instead. At last observation 18 people had progressed from an MCI diagnosis to a dementia diagnosis, giving a total of 22 dementia diagnoses (14 men, 8 women), with time to dementia diagnosis shown in Panel B.

**Figure 7:**
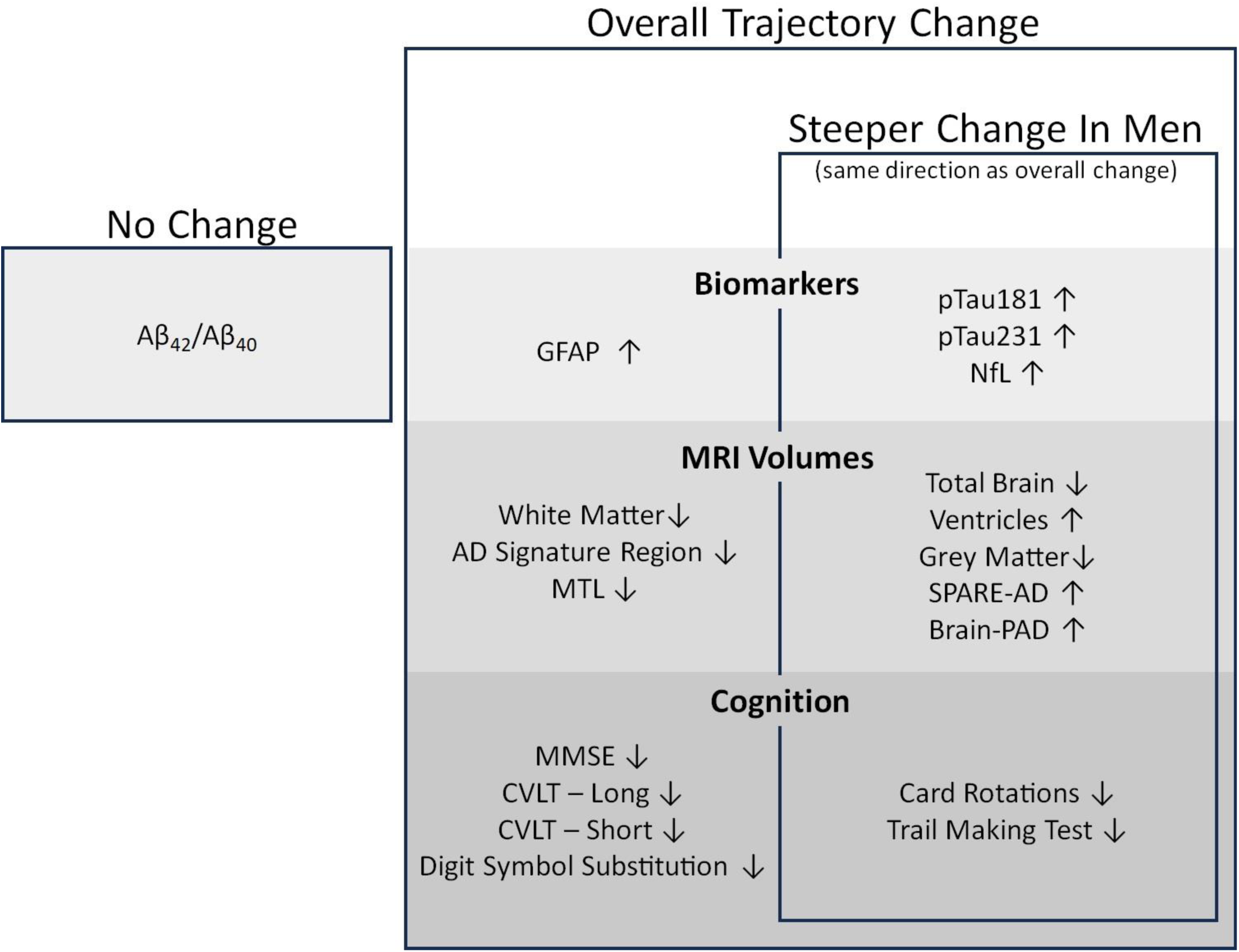
Summary of findings. In amyloid positive individuals, after centering on estimated age of amyloid onset, there are longitudinal changes in all investigated plasma biomarkers, MRI volumes, and cognitive outcomes except for Aβ_42_/Aβ_40_. The direction of this change is indicated by ↑ and ↓ to represent increases and decreases, respectively. In a subset of measures men experienced steeper rates of change as indicated by the sex × time interaction term; these measures can be found within the inner rectangle (“Steeper change in men”). For all those measures, these steeper rates of change for men were in a consistent direction to the overall rate of change.

## Discussion

Although sex differences in the incidence of AD dementia have been well documented [2, 33], whether sex influences the trajectory of AD pathology, neurodegenerative processes, and cognitive decline in the time that follows the onset of cortical amyloid-β remains largely unknown. The current study used longitudinal modeling to investigate trajectories of AD and neurodegenerative plasma biomarkers, MRI-defined brain volumes, and cognitive measures relative to age of amyloid onset as estimated using amyloid-β PET imaging. After estimated amyloid onset, we found longitudinal increases in markers of soluble pTau (pTau181 and pTau231), neuronal injury (NfL), and reactive astrogliosis (GFAP) at rates of approximately 0.06, 0.08, 0.07, and 0.06 standardized units per year, respectively. Relative to women, men had steeper increases in plasma measures of pTau181, pTau231, and NfL after amyloid-β onset. This suggests that after amyloid onset men may experience more rapid pathogenic tau phosphorylation and a more rapid uptick in neuronal injury compared to women. In support of the biomarker findings, a sex difference was also found for changes in brain volumes, with men showing greater rates of total and regional brain atrophy (including patterns of AD-like brain atrophy [SPARE-AD] and accelerated brain aging [Brain-PAD]) following amyloid-β onset. Measures of cognition and symptom progression also mirrored the biomarker findings, with men showing steeper declines than women in visuospatial and executive functioning following amyloid onset and a qualitatively shorter time from amyloid-β onset to diagnoses of cognitive impairment and dementia. Taken together, our findings demonstrate that after amyloid-β onset men experience greater acceleration of pathologic changes and symptomatic progression relative to women.

As demonstrated previously, we found increases in plasma-defined AD pathology (pTau), neuronal injury, and reactive astrogliosis, as well as decreases in brain volumes and cognition following amyloid-β onset [13, 34–36]. We extend these findings by additionally describing sex differences in the magnitudes of changes in these measures. The findings reported here are consistent with the results of previous studies from the BLSA which reported steeper declines in brain volume and on four cognitive measures among men [18]. Relatedly, Armstrong et al. found that amyloid status was predictive of volumetric declines in the ERC and PHG among amyloid-positive men, but not among amyloid-positive women [17]. Although the current study did not find sex differences in the MTL or AD Signature meta-ROIs (both of which contain the ERC and PHG) after amyloid-β onset, we did find differences in several metrics of global brain atrophy, including total brain volume. One implication of this finding is that men have an accelerated pTau-related neurodegeneration after amyloid-β onset that is not confined to brain regions or cognitive domains typically affected by AD pathology.

In support of our findings, one study conducted in the Alzheimer’s Disease Neuroimaging (ADNI) cohort reported that at the same level of plasma pTau181 women had higher levels of amyloid and tau deposition [37]. Put another way, at the same level of amyloid deposition (i.e., at the same time since amyloid onset) men had more plasma pTau181 than women. Another study reported that in those who have biomarker evidence for astrocyte reactivity (i.e., elevated GFAP levels) there is a larger cross-sectional association between amyloid-β PET levels and plasma pTau181, 217, and 231 in men relative to women [7]. These findings suggest astrocyte reactivity may account for the more rapid accumulation in pTau in men that we observed in the present study. We note, however, that not all studies have demonstrated findings that are consistent with our results. For example, a recent study in the ADNI cohort reported that after amyloid onset men had a 32% lower risk of developing cognitive impairment (defined as a Clinical Dementia Rating of 1 or higher), with a median survival time from estimated amyloid onset to impairment of 12.89 years for women versus 14.08 years for men [10]. Another study in the Harvard Aging Brain Study (HABS) and ADNI cohorts found higher levels of tau PET in women at the same level of amyloid PET, suggesting that tau burden may increase more quickly in women [38].

Although the exact cause of AD has yet to be fully established, in the standard amyloid cascade hypothesis model of AD pathogenesis, amyloid begins accumulating before the spread of tau from the entorhinal cortex to association cortices. Amyloid-β, and by some accounts, neuroinflammation, are proposed to be necessary for hyperphosphorylation of tau, subsequent neuronal injury, and cognitive decline [39–41]. Our study is one of very few studies to use estimated time since amyloid onset instead of age to examine the trajectory of brain volumes and cognitive performance [9, 14, 42]. As shown here, the time since amyloid onset provides a unique framework for investigating the progression of AD pathology. Benchmarking follow-up times based on amyloid onset enables the examination of biomarker changes and symptomatic progression relative to amyloid onset, better lining up the trajectories of the biomarkers by anchoring to a common biological event. Given that (i) the onset of amyloid accumulation is widely considered the initiating biological event in AD pathogenesis and (ii) the age of amyloid onset varies considerably from person to person, we believe that the examination of AD biology dynamics relative to amyloid onset (rather than age) may yield additional insight about the trajectory of distinct disease processes thought to be downstream of amyloid.

Recent clinical trial read outs have shown that reducing amyloid can slow accumulation of tau deposition and symptomatic progression [43–47]. In the context of our findings which show that men experience accelerated rates of pTau accumulation following amyloid-β onset and corresponding accelerated increases in neuronal injury, brain atrophy, and cognitive decline, as well as a shorter time to cognitive impairment and dementia, men may be expected to experience greater clinical benefit from amyloid clearing therapies. In fact, van Dyck et al. found that the benefit of lecanemab in minimizing decline on the clinical dementia rating sum of boxes (CDR-SB) was driven by men [43]. Our findings may also indicate that fluid biomarker thresholds used in clinical trial enrollment might need to account for the sex of the individual. Steeper ADRD biomarker increases, brain volume loss, and cognitive decline among men when centering these factors to a common biological event, rather than chronological age, suggests that men are more severely affected at lower levels of pathology. Thus one potential explanation for the higher prevalence of dementia among women is that men succumb to AD pathology more quickly [3, 33], while women are more resilient, experiencing slower decline and longer survival [48]. This is one explanation for a study which found that that cognitively normal women and women with MCI had higher levels of cortical amyloid and tau compared to men with a similar level of symptoms [49].

Given the mixed findings in literature, and unclear mechanisms through which sex may play a role in the manifestation of AD, sex remains an important biological factor to consider in future studies. Limitations of this study include a relatively small sample size, as we were restricted to only those in the BLSA who were recruited to the PET neuroimaging subsample and found to be positive for cortical amyloid-β. Additionally, the BLSA, although community based, is largely white and highly educated, so it is unclear whether these findings generalize to a more diverse cohort. We note additionally that since carriage of the *APOE*ε4 allele is associated with amyloid accumulation, carriers of that allele are overrepresented in this sample, however the percentage of carriers is equivalent between men and women. If our findings are replicated in other longitudinal cohorts, future studies will be needed to understand the biological or environmental drivers of the sex differences in AD pathogenesis after amyloid-β onset. Despite these limitations, we show consistently across multiple levels of measurement – biomarkers, brain volumes, and cognition – that after amyloid onset men experience steeper pathological changes and symptom progression. Our results shed light on the natural history of AD pathologic changes and the sex differences present in AD pathogenesis. They also point to the need for greater attention to sex differences in associations of fluid and imaging biomarkers with cognitive and brain outcomes.

## Data Availability

BLSA data are available upon request from https://www.blsa.nih.gov. All requests are reviewed by the BLSA Data Sharing Proposal Review Committee.

## Acknowledgements

Thank you to the Baltimore Longitudinal Study of Aging staff and participants.

## Conflicts of Interest

Henrik Zetterberg: HZ has served at scientific advisory boards and/or as a consultant for Abbvie, Acumen, Alector, Alzinova, ALZPath, Amylyx, Annexon, Apellis, Artery Therapeutics, AZTherapies, Cognito Therapeutics, CogRx, Denali, Eisai, LabCorp, Merry Life, Nervgen, Novo Nordisk, Optoceutics, Passage Bio, Pinteon Therapeutics, Prothena, Red Abbey Labs, reMYND, Roche, Samumed, Siemens Healthineers, Triplet Therapeutics, and Wave, has given lectures in symposia sponsored by Alzecure, Biogen, Cellectricon, Fujirebio, Lilly, Novo Nordisk, and Roche, and is a co-founder of Brain Biomarker Solutions in Gothenburg AB (BBS), which is a part of the GU Ventures Incubator Program (outside submitted work).

Keenan Walker: Associate Editor Alzheimer’s and Dementia, Associate Editor Alzheimer’s and Dementia: Translational Research & Clinical Interventions

Cassandra Joynes, Murat Bilgel, Yang An, Abhay R. Moghekar, Nicholas Ashton, Przemysław R. Kac, Thomas K. Karikari, Kaj Blennow, Madhav Thambisetty, Luigi Ferrucci, Susan M. Resnick: Nothing to disclose

## Consent Statement

This study was approved by the local institutional review boards overseeing the Baltimore Longitudinal Study of Aging and the PET imaging studies, and participants provided written informed consent at each study visit.

