## Supplemental Tables & Figures for "Sex differences in the trajectories of plasma biomarkers, brain atrophy, and cognitive decline relative to amyloid onset"

### Supplementary tables and figures

**sTable 1:** Demographic information for all those with amyloid PET imaging, both amyloid positive and negative

| | All | Amyloid – | Amyloid + |
|---|---|---|---|
| Number | 225 | 147 | 78 |
| APOE4 Carrier[a] | 65 (29.68%) | 36 (25.17%) | 29 (38.16%) |
| Sex (N Male) | 114 (50.67%) | 69 (46.93%) | 43 (56.58%) |
| *Race*[b] | | | |
| White | 175 (77.78%) | 111 (75.51%) | 64 (82.05%) |
| Non-White | 47 (61.84%) | 35 (46.05%) | 12 (15.79%) |
| *Cognitive Status (at end)* | | | |
| CN | 170 (75.56%) | 128 (87.07%) | 42 (53.85%) |
| MCI | 24 (10.67%) | 10 (6.8%) | 14 (17.95%) |
| Dementia | 31 (13.78%) | 9 (6.12%) | 22 (28.21%) |
| *Biomarker Measures* | | | |
| Age at First Measurement | 73.07 (8.03) | 71.92 (8.02) | 75.3 (7.61) |
| Number of Visits | 3.75 (2.07) | 3.6 (2.01) | 4.04 (2.18) |
| Follow up | | | |
| All measurements | 6.43 (3.94) | 6.26 (4.04) | 6.77 (3.74) |
| 2+ measurements | 7.11 (3.51) | 6.98 (3.62) | 7.35 (3.3) |

**Note:** The analysis from which these demographics were drawn was centered on each participant's first biomarker visit. Thus, the age at first measurement, number of visits, and follow up duration values are slightly longer here than those in Table 1, since some participants (n=13) had biomarker visits before they were amyloid positive, which were omitted in analyses which focused on only the timepoints after age of amyloid onset. [a] APOE status not available for 6 participants, [b] race information not available for 3 participants.

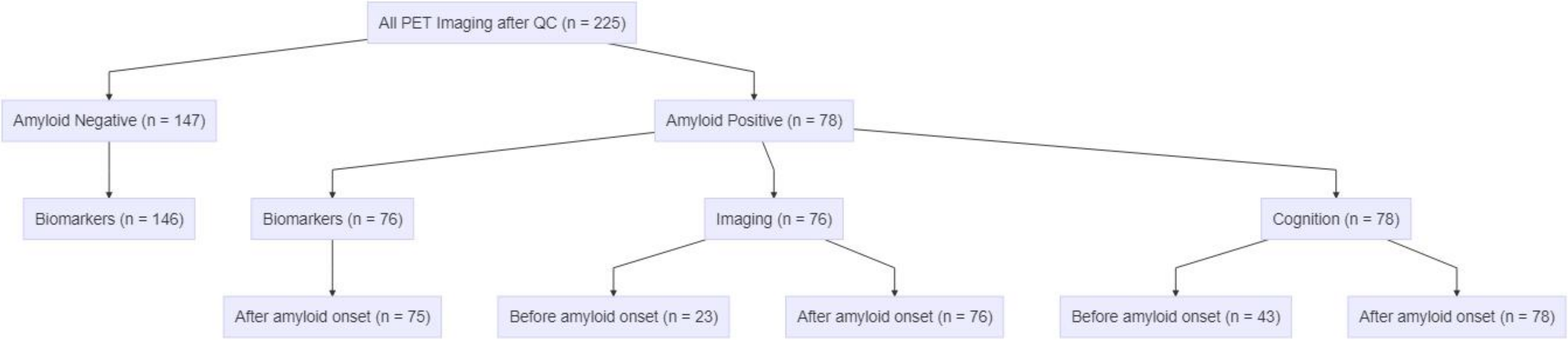


**sFigure 1: Consort diagram showing number of participants in each arm of analysis**. Before/after amyloid onset indicates the number of participants with data collected prior to/after their estimated age of amyloid onset. **Note:** the 76 with imaging measures are not the same 76 as those with biomarker measures.

**sTable 2**: Linear mixed effects models estimating plasma biomarker levels relative to age at plasma biomarker blood draw

| | Estimate (β) | Std. Error | p value |
|---|---|---|---|
| $A\beta_{42}/A\beta_{40}$ | | | |
| (Intercept) | **0.05** | **0.002** | **<2E-16** |
| Centered Age at First Plasma | -7.20E-05 | 7.76E-05 | 0.35 |
| Sex (M) | **-0.002** | **0.001** | **0.03** |
| eGFR | **6.68E-05** | **2.89E-05** | **0.02** |
| Amyloid Positive | **-0.008** | **0.001** | **3.13E-09** |
| Time Since First Plasma | **-3.88E-04** | **8.47E-05** | **5.58E-06** |
| Amyloid Positive × Time | **3.91E-04** | **1.44E-04** | **0.01** |
| *pTau181* | | | |
| (Intercept) | **10.09** | **1.77** | **1.98E-08** |
| Centered Age at First Plasma | **0.20** | **0.05** | **1.93E-04** |
| Sex (M) | **4.10** | **0.75** | **1.12E-07** |
| eGFR | **-0.07** | **0.02** | **0.002** |
| Amyloid Positive | 1.34 | 0.91 | 0.14 |
| Time Since First Plasma | -0.02 | 0.07 | 0.81 |
| Amyloid Positive × Time | **0.57** | **0.11** | **3.15E-07** |
| *pTau231* | | | |
| (Intercept) | **18.93** | **2.53** | **2.52E-13** |
| Centered Age at First Plasma | **0.21** | **0.08** | **0.01** |
| Sex (M) | **5.54** | **1.13** | **1.83E-06** |
| eGFR | **-0.11** | **0.03** | **1.37E-04** |
| Amyloid Positive | **6.20** | **1.34** | **5.61E-06** |
| Time Since First Plasma | **0.51** | **0.09** | **3.94E-08** |
| Amyloid Positive × Time | **0.63** | **0.15** | **4.05E-05** |
| *GFAP* | | | |
| (Intercept) | **223.45** | **21.22** | **<2E-16** |
| Centered Age at First Plasma | **4.09** | **0.72** | **4.02E-08** |
| Sex (M) | -13.22 | 10.74 | 0.22 |
| eGFR | **-0.98** | **0.24** | **5.96E-05** |
| Amyloid Positive | **40.65** | **12.03** | **8.34E-04** |
| Time Since First Plasma | **5.15** | **0.68** | **1.70E-13** |
| Amyloid Positive × Time | **2.96** | **1.15** | **0.01** |
| *NfL* | | | |
| (Intercept) | **35.43** | **2.96** | **<2E-16** |
| Centered Age at First Plasma | **0.57** | **0.08** | **7.66E-11** |
| Sex (M) | **2.98** | **1.22** | **0.02** |
| eGFR | **-0.24** | **0.03** | **1.10E-11** |
| Amyloid Positive | 1.48 | 1.45 | 0.31 |
| Time Since First Plasma | **0.93** | **0.11** | **<2E-16** |
| Amyloid Positive × Time | 0.32 | 0.18 | 0.08 |

Results derived from linear mixed-effects modeling of biomarker levels following first plasma biomarker draw, an amyloid status by time interaction term. Bold indicates significance at $p < 0.05$. All biomarkers except pTau181 show significant longitudinal increases. All except NfL show a significant interaction between time and amyloid positive status, indicating biomarker trajectories differ across those with those with detectable amyloid accumulation at any visit.
**Abbreviations:** GFAP – glial fibrillary acidic protein; NfL – neurofilament light; pTau – phosphorylated tau.

**sTable 3:** Demographic information for each subcategory of analysis in the amyloid positive subset

| | Full Sample | | | Pre-Onset | | | Post-Onset | | |
|---|---|---|---|---|---|---|---|---|---|
| | All | Males | Females | All | Males | Females | All | Males | Females |
| Biomarkers | | | | | | | | | |
| Number | -- | -- | -- | -- | -- | -- | 75 | 42 | 33 |
| APOE4 Carrier | -- | -- | -- | -- | -- | -- | 29 | 17 | 12 |
| *Race* | | | | | | | | | |
| White | -- | -- | -- | -- | -- | -- | 63 | 38 | 25 |
| Non-White | -- | -- | -- | -- | -- | -- | 12 | 4 | 8 |
| Estimated Age of Amyloid Onset | -- | -- | -- | -- | -- | -- | 68.16(9.1) | 68.8(9.44) | 67.34(8.71) |
| *Cognitive Status (at end)* | | | | | | | | | |
| CN | -- | -- | -- | -- | -- | -- | 40 | 21 | 19 |
| MCI | -- | -- | -- | -- | -- | -- | 13 | 7 | 6 |
| Dementia | -- | -- | -- | -- | -- | -- | 21 | 13 | 8 |
| MRI | | | | | | | | | |
| Number | 76 | 43 | 33 | 23 | 11 | 12 | 76 | 43 | 33 |
| APOE4 Carrier[a] | 28 | 16 | 12 | 7 | 1 | 6 | 28 | 16 | 12 |
| *Race*[b] | | | | | | | | | |
| White | 62 | 37 | 25 | 19 | 11 | 8 | 62 | 37 | 25 |
| Non-White | 12 | 4 | 8 | 4 | 0 | 4 | 12 | 4 | 8 |
| Estimated Age of Amyloid Onset | 68.01(9.07) | 68.52(9.4) | 67.34(8.71) | 72.2(7.23) | 74.23(6.91) | 70.33(7.3) | 68.01(9.07) | 68.52(9.4) | 67.34(8.71) |
| *Cognitive Status (at last visit)* | | | | | | | | | |
| CN | 40 | 21 | 19 | 11 | 4 | 7 | 40 | 21 | 19 |
| MCI | 13 | 7 | 6 | 5 | 2 | 3 | 13 | 7 | 6 |
| Dementia | 22 | 14 | 8 | 7 | 5 | 2 | 22 | 14 | 8 |
| Cognitive (MMSE) | | | | | | | | | |
| Number | 78 | 45 | 33 | 43 | 23 | 20 | 78 | 45 | 33 |
| APOE4 Carrier[a] | 29 | 17 | 12 | 13 | 6 | 7 | 29 | 17 | 12 |
| *Race*[b] | | | | | | | | | |
| White | 64 | 39 | 25 | 34 | 21 | 13 | 64 | 39 | 25 |
| Non-White | 12 | 4 | 8 | 9 | 2 | 7 | 12 | 4 | 8 |
| Estimated Age of Amyloid Onset | 68.11(9.01) | 68.68(9.28) | 67.34(8.71) | 71.45(7.56) | 72.28(7.8) | 70.5(7.36) | 68.11(9.01) | 68.68(9.28) | 67.34(8.71) |
| *Cognitive Status (at last visit)* | | | | | | | | | |
| CN | 41 | 22 | 19 | 24 | 11 | 13 | 41 | 22 | 19 |
| MCI | 14 | 8 | 6 | 8 | 4 | 4 | 14 | 8 | 6 |
| Dementia | 22 | 14 | 8 | 11 | 8 | 3 | 22 | 14 | 8 |

Values are provided as mean and standard deviation or counts with percentage of sample, as appropriate. NfL had one participant fewer than the other biomarkers. [a] APOE genotyping missing for 2 participants, [b] racial information missing for 2 participants.

**sTable 4:** Linear mixed effects models estimating plasma biomarker levels after estimated age of amyloid onset

| | Estimate (β) | Std. Error | p value | Estimate (β) | Std. Error | p value |
|---|---|---|---|---|---|---|
| $A\beta_{42}/A\beta_{40}$ | | | | | | |
| (Intercept) | 0.04 | 0.00 | <2E-16 | **0.04** | **0.00** | **<2E-16** |
| Years Since Amyloid Onset | -4.38E-05 | 1.00E-04 | 0.66 | -3.64E-05 | 1.52E-04 | 0.81 |
| Estimated Age of Amyloid Onset | -2.69E-05 | 1.09E-04 | 0.80 | -2.72E-05 | 1.09E-04 | 0.80 |
| Sex (M) | **-4.77E-03** | **1.65E-03** | **0.005** | -4.64E-03 | 2.63E-03 | 0.08 |
| eGFR | 8.29E-05 | 4.42E-05 | 0.06 | 8.31E-05 | 4.43E-05 | 0.06 |
| Sex × Time | -- | -- | -- | -1.13E-05 | 1.83E-04 | 0.95 |
| *pTau181* | | | | | | |
| (Intercept) | **8.95** | **2.91** | **0.0024** | **11.93** | **3.04** | **0.00011** |
| Years Since Amyloid Onset | **0.54** | **0.08** | **5.36E-11** | **0.26** | **0.12** | **0.03** |
| Estimated Age of Amyloid Onset | **0.39** | **0.10** | **9.62E-05** | **0.40** | **0.10** | **5.43E-05** |
| Sex (M) | **6.27** | **1.50** | **7.64E-05** | 1.16 | 2.26 | 0.61 |
| eGFR | **-0.08** | **0.03** | **0.017** | **-0.08** | **0.03** | **0.015** |
| Sex × Time | -- | -- | -- | **0.45** | **0.15** | **0.0025** |
| *pTau231* | | | | | | |
| (Intercept) | **30.11** | **5.02** | **7.85E-09** | **36.65** | **5.22** | **2.35E-11** |
| Years Since Amyloid Onset | **1.01** | **0.14** | **3.07E-12** | **0.44** | **0.21** | **0.036** |
| Estimated Age of Amyloid Onset | **0.42** | **0.16** | **0.0095** | **0.46** | **0.16** | **0.0051** |
| Sex (M) | **6.67** | **2.47** | **0.0087** | -3.91 | 3.81 | 0.31 |
| eGFR | **-0.27** | **0.06** | **7.09E-06** | **-0.27** | **0.06** | **2.62E-06** |
| Sex × Time | -- | -- | -- | **0.92** | **0.25** | **0.00031** |
| *GFAP* | | | | | | |
| (Intercept) | **296.33** | **43.13** | **4.51E-11** | **299.85** | **45.23** | **1.81E-10** |
| Years Since Amyloid Onset | **7.14** | **1.11** | **6.32E-10** | **6.79** | **1.72** | **9.52E-05** |
| Estimated Age of Amyloid Onset | **4.99** | **1.47** | **0.00096** | **5.01** | **1.47** | **0.00095** |
| Sex (M) | -13.95 | 23.81 | 0.56 | -20.08 | 33.30 | 0.55 |
| eGFR | **-1.68** | **0.49** | **0.00077** | **-1.68** | **0.50** | **0.00078** |
| Sex × Time | -- | -- | -- | 0.56 | 2.11 | 0.79 |
| *NfL* | | | | | | |
| (Intercept) | **40.53** | **6.32** | **9.69E-10** | **45.35** | **6.65** | **9.09E-11** |
| Years Since Amyloid Onset | **1.08** | **0.17** | **8.91E-10** | **0.62** | **0.25** | **0.015** |
| Estimated Age of Amyloid Onset | **0.74** | **0.17** | **3.93E-05** | **0.79** | **0.18** | **1.93E-05** |
| Sex (M) | 4.88 | 2.55 | 0.0598 | -3.77 | 4.27 | 0.38 |
| eGFR | **-0.36** | **0.07** | **3.01E-06** | **-0.36** | **0.07** | **2.97E-06** |
| Sex × Time | -- | -- | -- | **0.76** | **0.30** | **0.012** |

Results derived from linear mixed-effects modeling of biomarker levels after estimated age of amyloid onset, without (left) and with (right) a sex by time interaction term. Estimates for the sex and the sex × time interaction terms are relative to women. Thus, a positive β value for the sex term indicates that at baseline (amyloid onset) men have higher levels of the measure, whereas a positive β value for the interaction term indicates that men have more/steeper longitudinal increase in that metric than women. Bold indicates significance at $p < 0.05$. All biomarkers except $A\beta_{42}/A\beta_{40}$ show significant longitudinal increases. pTau181 and pTau231 have a significant interaction between time and sex, indicating pTau trajectories differ across sexes after amyloid onset. **Sample sizes:** GFAP – n observations = 290, n participants = 75; NfL – n observations: 286, n participants: 74; $A\beta_{42}/A\beta_{40}$ – n observations: 289, n participants: 75; pTau181 – n observations: 252, n participants: 75; pTau231– n observations: 253, n participants: 75. **Abbreviations:** Aβ – Amyloid β; GFAP – glial fibrillary acidic protein; NfL – neurofilament light; pTau – phosphorylated tau.

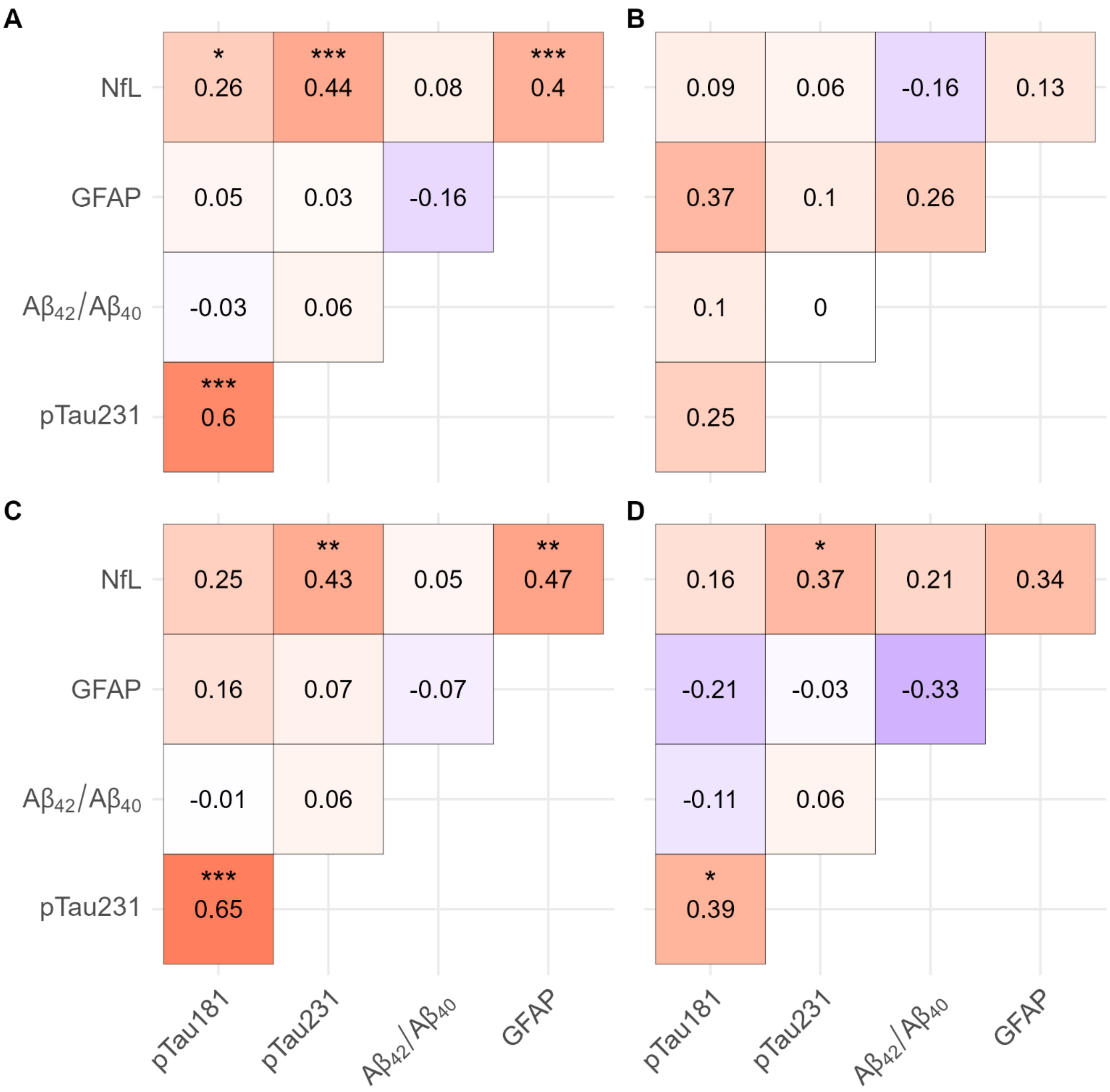


**sFigure 2: Relationships between longitudinal changes in biomarker slopes.** The Pearson correlations between individual slopes extracted from a linear mixed effects (LME) model. Unlike the primary linear effects models reported throughout the rest of the manuscript this LME model contained a random time term as well as a random intercept term, allowing for the extraction of each participant's individualized slope. In all panels colors and numbers indicate the strength and direction of the correlation (red = positive; blue = negative) while asterisks indicate the significance (* = $p < 0.05$; ** = $p < 0.01$; *** = $p < 0.001$) **A.** Pearson correlations between pairs of two biomarkers for all 75 participants with biomarker measurements following amyloid onset **B.** The difference in correlation size (and direction) between subsets of male and female participants only. Difference is calculated as male minus female meaning red indicates a more positive correlation for males. Significance levels are not reported for **B** as it shows a subtraction of two other correlation values. **C-D.** Pearson correlations between longitudinal biomarker slopes in only males **(C)** and only females **(D)**. **Abbreviations:** GFAP – glial fibrillary acidic protein; NfL – neurofilament light chain; pTau – phosphorylated tau

**sTable 5:** Linear mixed effects models estimating MRI-defined brain volumes and MRI derived brain health measures pre- and post- estimated age of amyloid onset without sex by time interaction term

| | Pre Onset | | | Post Onset | | |
|---|---|---|---|---|---|---|
| | Estimate (β) | Std. Error | p value | Estimate (β) | Std. Error | p value |
| *Total Brain Volume* | | | | | | |
| (Intercept) | -15564.35 | 11627.70 | 0.20 | 3045.85 | 6565.66 | 0.64 |
| Years Since Amyloid Onset | **-6501.95** | **392.44** | **5.76E-26** | **-5610.00** | **122.70** | **3.75E-163** |
| Estimated Age of Amyloid Onset | **-4375.86** | **1195.45** | **0.002** | **-5250.02** | **475.59** | **2.47E-17** |
| Sex (M) | -705.36 | 16932.82 | 0.97 | 8151.95 | 8610.96 | 0.35 |
| *Grey Matter* | | | | | | |
| (Intercept) | -4667.24 | 5426.89 | 0.40 | **10262.91** | **3934.68** | **0.01** |
| Years Since Amyloid Onset | **-5160.96** | **272.63** | **7.88E-30** | **-4833.67** | **92.22** | **3.27E-186** |
| Estimated Age of Amyloid Onset | **-3090.27** | **553.82** | **1.75E-05** | **-3903.42** | **283.29** | **3.25E-22** |
| Sex (M) | -1977.63 | 7850.96 | 0.80 | 5829.18 | 5117.33 | 0.26 |
| *White Matter* | | | | | | |
| (Intercept) | -12345.01 | 7415.17 | 0.11 | -4597.11 | 4041.79 | 0.26 |
| Years Since Amyloid Onset | **-2134.46** | **172.07** | **3.16E-19** | **-2109.32** | **53.29** | **8.57E-141** |
| Estimated Age of Amyloid Onset | **-1722.30** | **764.76** | **0.04** | **-2284.32** | **294.29** | **3.71E-11** |
| Sex (M) | 1159.75 | 10828.36 | 0.92 | 1679.62 | 5338.93 | 0.75 |
| *Ventricular Volume* | | | | | | |
| *(Intercept)* | 1495.48 | 3488.92 | 0.67 | -2254.70 | 3055.37 | 0.46 |
| Years Since Amyloid Onset | **923.78** | **63.44** | **1.15E-22** | **1402.29** | **38.75** | **1.70E-128** |
| Estimated Age of Amyloid Onset | 671.51 | 360.22 | 0.08 | **1024.45** | **222.55** | **1.70E-05** |
| Sex (M) | -1310.41 | 5099.71 | 0.80 | 311.53 | 4038.05 | 0.94 |
| *Medial Temporal Lobe*[a] | | | | | | |
| (Intercept) | 414.49 | 469.93 | 0.39 | 241.50 | 212.42 | 0.26 |
| Years Since Amyloid Onset | **-118.21** | **13.24** | **3.82E-13** | **-100.41** | **4.10** | **1.82E-82** |
| Estimated Age of Amyloid Onset | **-113.00** | **48.40** | **0.03** | **-88.31** | **15.38** | **1.89E-07** |
| Sex (M) | 773.34 | 685.43 | 0.27 | **609.61** | **278.33** | **0.03** |
| *AD Signature Region*[b] | | | | | | |
| (Intercept) | 283.35 | 711.70 | 0.69 | 682.38 | 441.00 | 0.13 |
| Years Since Amyloid Onset | **-305.69** | **24.87** | **3.61E-19** | **-281.63** | **7.71** | **1.08E-130** |
| Estimated Age of Amyloid Onset | **-164.83** | **73.14** | **0.04** | **-222.27** | **31.99** | **1.20E-09** |
| Sex (M) | 372.61 | 1036.01 | 0.72 | 650.17 | 579.41 | 0.27 |
| *SPARE-AD* | | | | | | |
| (Intercept) | **-1.68** | **0.22** | **1.35E-07** | **-1.92** | **0.14** | **2.82E-22** |
| Years Since Amyloid Onset | **0.04** | **0.01** | **0.002** | **0.10** | **0.00** | **4.26E-96** |
| Estimated Age of Amyloid Onset | 0.04 | 0.02 | 0.07 | **0.07** | **0.01** | **5.62E-10** |
| Sex (M) | 0.27 | 0.31 | 0.39 | 0.03 | 0.19 | 0.87 |
| *Brain-PAD*[c] | | | | | | |
| (Intercept) | 1.96 | 1.52 | 0.21 | -1.54 | 1.18 | 0.20 |
| Years Since Amyloid Onset | **0.49** | **0.08** | **6.85E-08** | **0.19** | **0.03** | **3.94E-10** |
| Estimated Age of Amyloid Onset | **-0.46** | **0.16** | **0.009** | -0.11 | 0.08 | 0.22 |
| Sex (M) | 3.49 | 2.05 | 0.11 | -0.98 | 1.53 | 0.52 |

Results derived from linear mixed-effects modeling of MRI volume measures before and after estimated age of amyloid onset. Estimates for the sex term is relative to women; thus, a negative β value for the sex term indicates that at baseline (amyloid onset, for the post-onset LME) men have lower levels of the measure. Bold indicates significance at $p < 0.05$.

**Sample sizes (pre|post):** n observations: 92|471, n participants: 23|76. **Notes:** [a] MTL consists of hippocampus, entorhinal cortex, parahippocampal gyrus, and amygdala [b] AD Signature region consists of hippocampus, entorhinal cortex, parahippocampal gyrus, precuneus, and posterior cingulate gyrus [c] Brain-PAD is calculated as SPARE-Brain Age minus chronological age for each MRI. **Abbreviations:** AD – Alzheimer's disease; Brain-PAD – Brain predicted age difference; MTL – medial temporal lobe; SPARE-AD – Spatial Pattern of Abnormality for Recognition of Early Alzheimer's Disease.

**sTable 6:** Linear mixed effects models estimating MRI-defined brain volumes and MRI derived brain health measures pre- and post- estimated age of amyloid onset including sex by time interaction term

| | Pre Onset | | | Post Onset | | |
|---|---|---|---|---|---|---|
| | Estimate (β) | Std. Error | p value | Estimate (β) | Std. Error | p value |
| *Total Brain Volume* | | | | | | |
| (Intercept) | -6593.52 | 11905.78 | 0.59 | -1715.60 | 6784.18 | 0.80 |
| Years Since Amyloid Onset | **-3655.86** | **691.73** | **1.40E-06** | **-5067.12** | **219.55** | **2.73E-76** |
| Estimated Age of Amyloid Onset | **-4620.84** | **1211.54** | **0.001** | **-5254.81** | **477.64** | **2.95E-17** |
| Sex (M) | -11478.94 | 17292.56 | 0.51 | 15346.62 | 8982.29 | 0.09 |
| Sex × Time | **-3798.88** | **795.61** | **9.81E-06** | **-779.91** | **262.75** | **0.003** |
| *Grey Matter* | | | | | | |
| (Intercept) | 1425.91 | 5622.92 | 0.80 | 5765.76 | 4127.32 | 0.17 |
| Years Since Amyloid Onset | **-3235.03** | **485.73** | **4.85E-09** | **-4322.67** | **163.75** | **1.91E-91** |
| Estimated Age of Amyloid Onset | **-3254.10** | **560.45** | **1.02E-05** | **-3907.80** | **284.55** | **4.06E-22** |
| Sex (M) | -9345.90 | 8088.22 | 0.26 | **12624.92** | **5450.62** | **0.02** |
| Sex × Time | **-2573.41** | **559.21** | **1.79E-05** | **-734.84** | **195.76** | **2E-04** |
| *White Matter* | | | | | | |
| (Intercept) | -8598.77 | 7431.32 | 0.26 | -5966.47 | 4101.09 | 0.15 |
| Years Since Amyloid Onset | **-938.99** | **309.04** | **0.003** | **-1952.71** | **96.14** | **1.11E-63** |
| Estimated Age of Amyloid Onset | **-1821.63** | **762.29** | **0.03** | **-2285.65** | **294.23** | **3.61E-11** |
| Sex (M) | -3353.29 | 10835.17 | 0.76 | 3751.55 | 5442.14 | 0.49 |
| Sex × Time | **-1587.69** | **355.32** | **3.07E-05** | -225.01 | 115.16 | 0.05 |
| *Ventricular Volume* | | | | | | |
| *(Intercept)* | 454.56 | 3432.70 | 0.90 | -9.77 | 3081.31 | 0.997 |
| Years Since Amyloid Onset | **589.89** | **121.64** | **7.60E-06** | **1145.50** | **68.57** | **5.77E-48** |
| Estimated Age of Amyloid Onset | 698.48 | 352.92 | 0.06 | **1026.59** | **221.51** | **1.51E-05** |
| Sex (M) | -55.59 | 5010.53 | 0.99 | -3087.81 | 4089.87 | 0.45 |
| Sex × Time | **440.96** | **139.84** | **0.002** | **369.11** | **82.14** | **9.16E-06** |
| *Medial Temporal Lobe*[a] | | | | | | |
| (Intercept) | 655.80 | 478.25 | 0.18 | 251.67 | 219.18 | 0.25 |
| Years Since Amyloid Onset | -40.65 | 24.68 | 0.10 | **-101.56** | **7.41** | **1.63E-35** |
| Estimated Age of Amyloid Onset | **-119.46** | **48.84** | **0.02** | **-88.30** | **15.37** | **1.89E-07** |
| Sex (M) | 482.86 | 695.80 | 0.50 | **594.25** | **290.06** | **0.04** |
| Sex × Time | **-103.14** | **28.38** | **5E-04** | 1.66 | 8.87 | 0.85 |
| *AD Signature Region*[b] | | | | | | |
| (Intercept) | 862.47 | 712.62 | 0.24 | 495.53 | 453.41 | 0.28 |
| Years Since Amyloid Onset | **-120.51** | **43.78** | **0.008** | **-260.31** | **13.90** | **5.28E-57** |
| Estimated Age of Amyloid Onset | **-180.22** | **72.37** | **0.02** | **-222.45** | **32.05** | **1.25E-09** |
| Sex (M) | -330.53 | 1034.08 | 0.75 | 932.70 | 600.62 | 0.12 |
| Sex × Time | **-246.38** | **50.36** | **6.30E-06** | -30.63 | 16.64 | 0.07 |
| *SPARE-AD* | | | | | | |
| (Intercept) | **-1.77** | **0.23** | **4.81E-08** | **-1.67** | **0.15** | **2.19E-18** |
| Years Since Amyloid Onset | 0.01 | 0.02 | 0.71 | **0.07** | **0.01** | **3.04E-26** |
| Estimated Age of Amyloid Onset | 0.04 | 0.02 | 0.06 | **0.07** | **0.01** | **1.00E-09** |
| Sex (M) | 0.38 | 0.32 | 0.26 | -0.36 | 0.20 | 0.08 |
| Sex × Time | 0.04 | 0.03 | 0.15 | **0.04** | **0.01** | **1.78E-07** |
| *Brain-PAD*[c] | | | | | | |

| | | | | | | |
|---|---|---|---|---|---|---|
| (Intercept) | 0.44 | 1.57 | 0.78 | -0.19 | 1.26 | 0.88 |
| Years Since Amyloid Onset | 0.02 | 0.16 | 0.92 | 0.04 | 0.05 | 0.46 |
| Estimated Age of Amyloid Onset | **-0.42** | **0.15** | **0.01** | -0.10 | 0.09 | 0.23 |
| Sex (M) | **5.29** | **2.11** | **0.02** | -2.99 | 1.66 | 0.08 |
| Sex × Time | **0.61** | **0.19** | **0.002** | **0.22** | **0.06** | **7E-04** |

Results derived from linear mixed-effects modeling of MRI volume measures before and after estimated age of amyloid onset. Estimates for the sex and the sex × time interaction terms are relative to women. Thus, a negative β value for the sex term indicates that at baseline (amyloid onset) men have lower levels of the measure, whereas a negative β value for the interaction term indicates that men have more/steeper longitudinal decline in that metric than women. Bold indicates significance at $p < 0.05$. **Sample sizes (pre|post):** n observations: 92|471, n participants: 23|76. **Notes:** [a] MTL consists of hippocampus, entorhinal cortex, parahippocampal gyrus, and amygdala [b] AD Signature region consists of hippocampus, entorhinal cortex, parahippocampal gyrus, precuneus, and posterior cingulate gyrus [c] Brain-PAD is calculated as SPARE-Brain Age minus chronological age for each MRI. **Abbreviations:** AD – Alzheimer's disease; Brain-PAD – Brain predicted age difference; MTL – medial temporal lobe; SPARE-AD – Spatial Pattern of Abnormality for Recognition of Early Alzheimer's Disease.

**sTable 7:** Linear mixed effects models of cognitive measures pre- and post- estimated age of amyloid onset without the sex by time interaction term

| | Pre Onset | | | Post Onset | | |
|---|---|---|---|---|---|---|
| | Estimate (β) | Std. Error | p value | Estimate (β) | Std. Error | p value |
| *MMSE* | | | | | | |
| (Intercept) | **29.24** | **0.25** | **3.80E-73** | **29.85** | **0.24** | **7.05E-132** |
| Years Since Amyloid Onset | -0.02 | 0.02 | 0.26 | **-0.13** | **0.01** | **8.41E-30** |
| Estimated Age of Amyloid Onset | -0.03 | 0.02 | 0.10 | **-0.08** | **0.02** | **6.82E-07** |
| Sex (M) | -0.31 | 0.31 | 0.32 | -0.33 | 0.27 | 0.23 |
| *Trails A Minus Trails B* | | | | | | |
| (Intercept) | **-37.42** | **4.47** | **1.45E-11** | **-34.64** | **5.33** | **6.62E-09** |
| Years Since Amyloid Onset | -0.34 | 0.41 | 0.41 | **-2.34** | **0.21** | **2.90E-26** |
| Estimated Age of Amyloid Onset | -0.53 | 0.37 | 0.16 | **-1.61** | **0.36** | **3.61E-05** |
| Sex (M) | -3.54 | 5.44 | 0.52 | -2.15 | 6.48 | 0.74 |
| *CVLT - List Learning* | | | | | | |
| (Intercept) | **59.85** | **1.98** | **1.61E-32** | **63.25** | **1.99** | **7.83E-52** |
| Years Since Amyloid Onset | 0.04 | 0.11 | 0.70 | **-1.05** | **0.06** | **2.15E-50** |
| Estimated Age of Amyloid Onset | -0.08 | 0.16 | 0.59 | **-0.67** | **0.14** | **6.08E-06** |
| Sex (M) | **-6.92** | **2.75** | **0.02** | **-5.52** | **2.50** | **0.03** |
| *CVLT - Long Recall* | | | | | | |
| (Intercept) | **13.12** | **0.53** | **7.15E-29** | **14.00** | **0.57** | **3.03E-42** |
| Years Since Amyloid Onset | 0.03 | 0.03 | 0.34 | **-0.30** | **0.02** | **5.17E-52** |
| Estimated Age of Amyloid Onset | -0.03 | 0.04 | 0.42 | **-0.20** | **0.04** | **4.31E-06** |
| Sex (M) | **-1.53** | **0.74** | **0.04** | -1.15 | 0.71 | 0.11 |
| *Digit Symbol Substitution* | | | | | | |
| (Intercept) | **45.69** | **3.01** | **3.50E-11** | **54.46** | **1.91** | **4.20E-47** |
| Years Since Amyloid Onset | **-1.39** | **0.44** | **0.007** | **-1.26** | **0.07** | **1.75E-56** |
| Estimated Age of Amyloid Onset | 0.11 | 0.38 | 0.77 | **-0.76** | **0.13** | **1.53E-07** |
| Sex (M) | -0.10 | 5.04 | 0.98 | -1.59 | 2.36 | 0.50 |
| *Card Rotations* | | | | | | |
| (Intercept) | **91.71** | **6.73** | **7.37E-18** | **88.51** | **6.01** | **2.06E-24** |
| Years Since Amyloid Onset | 0.30 | 0.33 | 0.37 | **-1.42** | **0.12** | **4.04E-31** |
| Estimated Age of Amyloid Onset | **-1.54** | **0.54** | **0.007** | **-1.51** | **0.43** | **8E-04** |
| Sex (M) | **23.93** | **9.48** | **0.02** | **17.13** | **7.81** | **0.03** |

Results derived from linear mixed-effects modeling of cognitive measures prior to and after estimated age of amyloid onset. Estimates for the sex term are relative to women; thus, a negative β value for the sex term indicates that at baseline (amyloid onset, for the post-onset LME) men have lower levels of the measure. Bold indicates significance at $p < 0.05$. **Sample sizes (pre|post):** CVLT list learning – n observations: 198|653, n participants: 43|77; CVLT long delay – n observations: 198|649, n participants: 43|77; MMSE – n observations: 195|702, n participants: 43|78; Trails B minus A – n observations: 192|668, n participants: 42|78; Digit Symbol – n observations: 28|386, n participants: 15|76; Card Rotations – n observations: 200|660, n participants: 43|77. **Abbreviations:** CVLT – California Verbal Learning Test, MMSE – Mini Mental State Examination.

**sTable 8:** Linear mixed effects models of cognitive measures pre- and post- estimated age of amyloid onset including sex by time interaction term

| | Pre Onset | | | Post Onset | | |
|---|---|---|---|---|---|---|
| | Estimate (β) | Std. Error | p value | Estimate (β) | Std. Error | p value |
| *MMSE* | | | | | | |
| (Intercept) | **29.27** | **0.29** | **2.89E-100** | **29.63** | **0.28** | **1.87E-184** |
| Years Since Amyloid Onset | -0.02 | 0.04 | 0.70 | **-0.11** | **0.02** | **1.52E-08** |
| Estimated Age of Amyloid Onset | -0.04 | 0.02 | 0.10 | **-0.08** | **0.02** | **6.51E-07** |
| Sex (M) | -0.36 | 0.37 | 0.33 | 0.02 | 0.36 | 0.96 |
| Sex × Time | -0.01 | 0.05 | 0.81 | -0.03 | 0.02 | 0.14 |
| *Trails A Minus Trails B* | | | | | | |
| (Intercept) | **-43.11** | **5.22** | **8.10E-13** | **-39.98** | **5.98** | **8.10E-10** |
| Years Since Amyloid Onset | **-1.63** | **0.74** | **0.03** | **-1.77** | **0.36** | **8.57E-07** |
| Estimated Age of Amyloid Onset | -0.47 | 0.37 | 0.21 | **-1.62** | **0.36** | **3.65E-05** |
| Sex (M) | 4.27 | 6.58 | 0.52 | 6.05 | 7.72 | 0.43 |
| Sex × Time | **1.85** | **0.88** | **0.04** | **-0.86** | **0.43** | **0.048** |
| *CVLT - List Learning* | | | | | | |
| (Intercept) | **60.18** | **2.05** | **5.72E-35** | **64.55** | **2.15** | **5.84E-59** |
| Years Since Amyloid Onset | 0.10 | 0.15 | 0.50 | **-1.19** | **0.11** | **1.28E-25** |
| Estimated Age of Amyloid Onset | -0.09 | 0.16 | 0.57 | **-0.67** | **0.14** | **6.52E-06** |
| Sex (M) | **-7.55** | **2.94** | **0.01** | **-7.55** | **2.81** | **0.008** |
| Sex × Time | -0.14 | 0.23 | 0.55 | 0.21 | 0.13 | 0.12 |
| *CVLT - Long Recall* | | | | | | |
| (Intercept) | **13.01** | **0.55** | **2.00E-30** | **14.34** | **0.61** | **6.94E-47** |
| Years Since Amyloid Onset | 0.01 | 0.04 | 0.81 | **-0.34** | **0.03** | **5.65E-26** |
| Estimated Age of Amyloid Onset | -0.03 | 0.04 | 0.45 | **-0.20** | **0.04** | **4.62E-06** |
| Sex (M) | -1.31 | 0.79 | 0.10 | **-1.67** | **0.80** | **0.04** |
| Sex × Time | 0.05 | 0.06 | 0.44 | 0.05 | 0.04 | 0.15 |
| *Digit Symbol Substitution* | | | | | | |
| (Intercept) | **45.34** | **3.01** | **2.05E-11** | **54.28** | **2.03** | **1.99E-50** |
| Years Since Amyloid Onset | **-1.51** | **0.49** | **0.009** | **-1.25** | **0.09** | **7.08E-34** |
| Estimated Age of Amyloid Onset | 0.08 | 0.37 | 0.83 | **-0.76** | **0.13** | **1.50E-07** |
| Sex (M) | 1.96 | 5.72 | 0.74 | -1.20 | 2.83 | 0.67 |
| Sex × Time | 0.99 | 1.39 | 0.49 | -0.03 | 0.13 | 0.80 |
| *Card Rotations* | | | | | | |
| (Intercept) | **92.81** | **6.86** | **1.52E-18** | **84.03** | **6.10** | **9.00E-24** |
| Years Since Amyloid Onset | 0.50 | 0.43 | 0.26 | **-0.92** | **0.20** | **4.97E-06** |
| Estimated Age of Amyloid Onset | **-1.56** | **0.54** | **0.006** | **-1.51** | **0.43** | **7E-04** |
| Sex (M) | **21.75** | **9.89** | **0.03** | **24.13** | **8.03** | **0.003** |
| Sex × Time | -0.49 | 0.68 | 0.47 | **-0.75** | **0.24** | **0.002** |

Results derived from linear mixed-effects modeling of cognitive measures prior to and after estimated age of amyloid onset. Estimates for the sex and the sex × time interaction terms are relative to women. Thus, a negative β value for the sex term indicates that at baseline (amyloid onset, for the post-onset LME) men have lower levels of the measure, whereas a negative β value for the interaction term indicates that men have more/steeper longitudinal decline in that metric than women. Bold indicates significance at $p < 0.05$. **Sample sizes (pre|post):** CVLT list learning – n observations: 198|653, n participants: 43|77; CVLT long delay – n observations: 198|649, n participants: 43|77; MMSE – n observations: 195|702, n participants: 43|78; Trails B minus A – n observations: 192|668, n participants: 42|78; Digit Symbol – n observations: 28|386, n participants: 15|76; Card Rotations – n observations: 200|660, n participants: 43|77. **Abbreviations:** CVLT – California Verbal Learning Test, MMSE – Mini Mental State Examination.

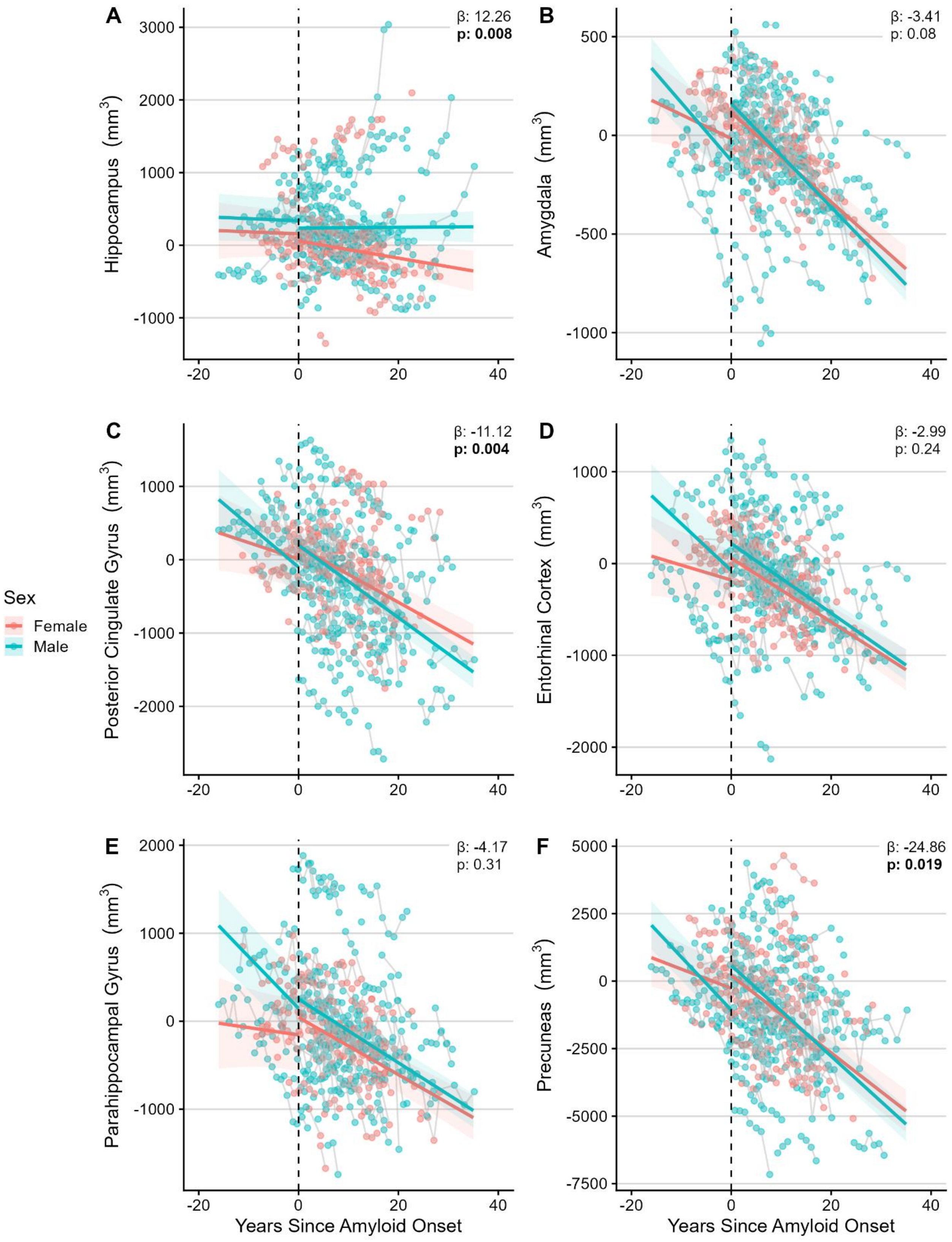


**sFigure 3: Sex-specific trajectories of brain volumes before and after estimated age of amyloid onset.** Sex specific trajectories of MRI-defined brain volume as estimated by linear mixed effects models for brain regions which comprised the composite AD Signature and MTL regions. β estimates and p-values listed on each panel are for the post-onset sex × time interaction term in each linear mixed effect model. Estimates for the interaction term are relative to women. Thus, a

negative β value for the interaction term indicates that men have more/steeper longitudinal decline in that metric than women. **Abbreviations:** AD – Alzheimer's Disease; MTL – Medial Temporal Lobe

**sTable 9:** Linear mixed effects models for MRI derived volumes of the brain regions included in the AD Signature and MTL composite regions

| | Pre Onset | | | Post Onset | | |
|---|---|---|---|---|---|---|
| | Estimate (β) | Std. Error | p value | Estimate (β) | Std. Error | p value |
| *Hippocampus* | | | | | | |
| (Intercept) | 135.89 | 151.65 | 0.38 | 56.88 | 106.05 | 0.59 |
| Years Since Amyloid Onset | -2.72 | 7.82 | 0.73 | **-11.78** | **3.81** | **0.002** |
| Estimated Age of Amyloid Onset | 2.98 | 15.49 | 0.85 | -4.18 | 7.40 | 0.57 |
| Sex (M) | 178.61 | 220.63 | 0.43 | 178.94 | 140.26 | 0.21 |
| Sex × Time | -0.33 | 8.99 | 0.97 | **12.28** | **4.55** | **0.007** |
| *Amygdala* | | | | | | |
| (Intercept) | 131.58 | 66.68 | 0.06 | **110.78** | **40.17** | **0.007** |
| Years Since Amyloid Onset | -12.16 | 5.99 | 0.05 | **-22.64** | **1.63** | **2.64E-36** |
| Estimated Age of Amyloid Onset | **-20.55** | **6.62** | **0.006** | **-18.83** | **2.76** | **2.01E-09** |
| Sex (M) | -111.62 | 95.77 | 0.26 | 45.59 | 53.03 | 0.39 |
| Sex × Time | **-17.21** | **6.90** | **0.01** | -3.59 | 1.95 | 0.07 |
| *Posterior Cingulate Gyrus* | | | | | | |
| (Intercept) | 37.70 | 190.14 | 0.84 | 172.07 | 115.49 | 0.14 |
| Years Since Amyloid Onset | -22.50 | 11.68 | 0.06 | **-37.95** | **3.16** | **1.03E-28** |
| Estimated Age of Amyloid Onset | -4.09 | 19.31 | 0.83 | **-26.14** | **8.23** | **0.002** |
| Sex (M) | -110.15 | 275.91 | 0.69 | 20.34 | 153.12 | 0.89 |
| Sex × Time | **-35.17** | **13.44** | **0.01** | **-11.50** | **3.78** | **0.002** |
| *Entorhinal Cortex* | | | | | | |
| (Intercept) | 208.57 | 155.72 | 0.19 | 50.69 | 85.12 | 0.55 |
| Years Since Amyloid Onset | -16.30 | 10.67 | 0.13 | **-34.75** | **3.06** | **2.90E-26** |
| Estimated Age of Amyloid Onset | **-53.94** | **15.74** | **0.003** | **-33.26** | **5.94** | **3.37E-07** |
| Sex (M) | 103.93 | 225.47 | 0.65 | 158.60 | 112.57 | 0.16 |
| Sex × Time | **-34.69** | **12.27** | **0.006** | -3.09 | 3.66 | 0.40 |
| *Parahippocampal Gyrus* | | | | | | |
| (Intercept) | 187.00 | 190.60 | 0.34 | 35.90 | 97.29 | 0.71 |
| Years Since Amyloid Onset | -8.32 | 12.18 | 0.50 | **-32.56** | **3.42** | **1.36E-19** |
| Estimated Age of Amyloid Onset | **-47.58** | **19.33** | **0.02** | **-31.91** | **6.80** | **1.20E-05** |
| Sex (M) | 301.78 | 276.37 | 0.29 | 207.73 | 128.70 | 0.11 |
| Sex × Time | **-50.44** | **14.02** | **6E-04** | -3.63 | 4.09 | 0.37 |
| *Precuneus* | | | | | | |
| (Intercept) | 293.25 | 418.98 | 0.49 | 180.27 | 337.33 | 0.59 |
| Years Since Amyloid Onset | **-71.15** | **22.36** | **0.002** | **-143.41** | **8.69** | **3.79E-47** |
| Estimated Age of Amyloid Onset | -77.57 | 42.75 | 0.08 | **-126.88** | **24.10** | **1.33E-06** |
| Sex (M) | -808.14 | 609.31 | 0.20 | 369.20 | 447.42 | 0.41 |
| Sex × Time | **-125.55** | **25.72** | **6.63E-06** | **-24.61** | **10.41** | **0.02** |

Results derived from linear mixed-effects modeling of MRI volume measures before and after estimated age of amyloid onset. Estimates for the sex and the sex × time interaction terms are relative to women. Thus, a negative β value for the sex term indicates that at baseline (amyloid onset) men have lower levels of the measure, whereas a negative β value for the interaction term indicates that men have more/steeper longitudinal decline in that metric than women. Bold indicates significance at $p < 0.05$. Individual LMEs were calculated for each region used in the composite AD Signature and MTL ROIs.

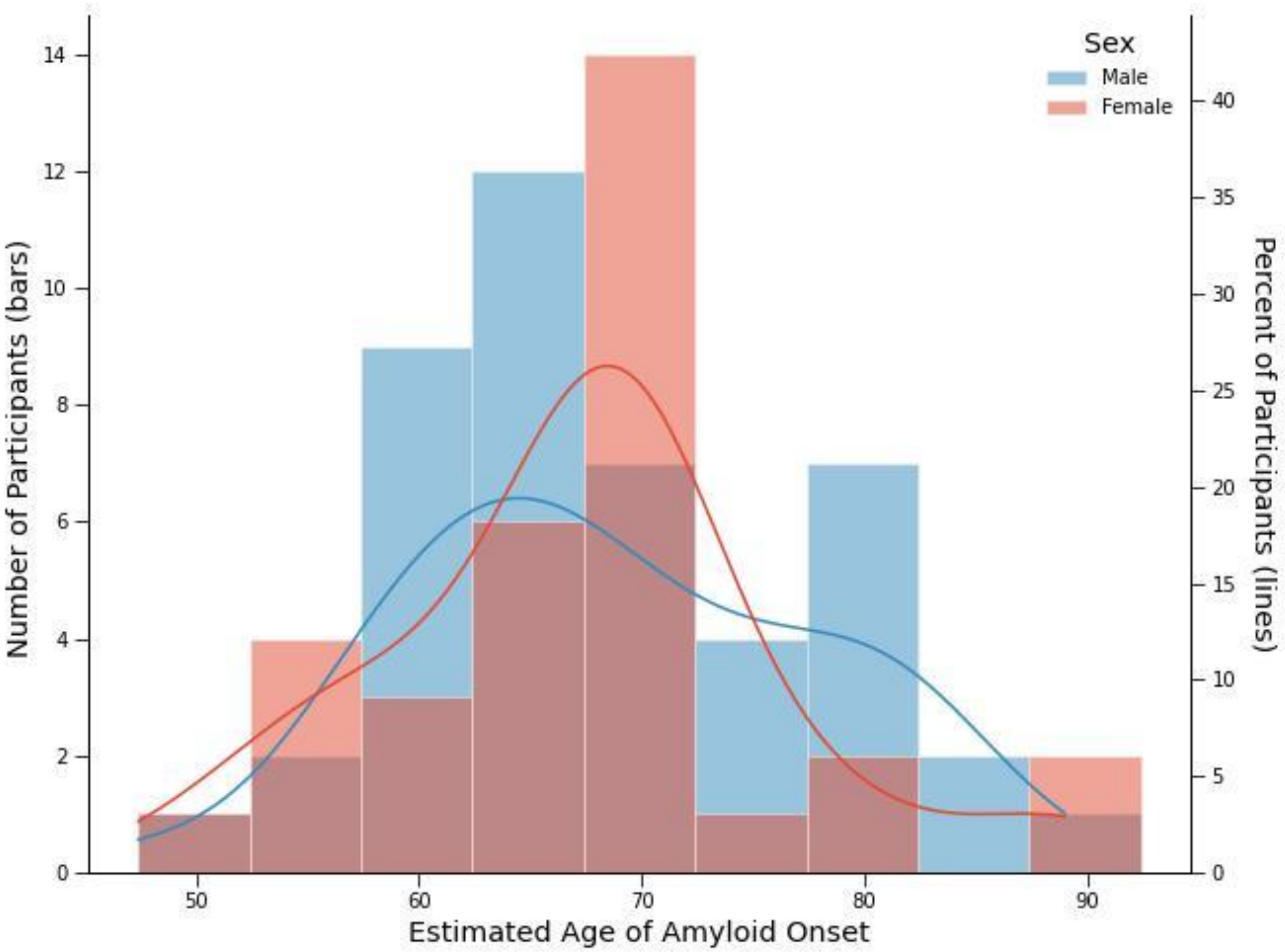


**sFigure 4:** Distribution of estimated age of amyloid onset, stratified by sex. Males had a mean estimated age of onset of 68.68 years, and females had a mean estimated age of onset of 67.34 years; see **Table 1** for additional details.
